# Natural-language cohort queries on a hospital data platform: accuracy and provenance in a real-world evaluation

**DOI:** 10.64898/2026.11.09.26367645

**Authors:** Dmitriy Desser, Peter Brunecker, Andrea Dell’Orco, Gabi Galperin, Mike P. Wattjes, Jawed Nawabi

## Abstract

**Background:** Accessing routinely collected clinical data for research remains technically demanding across heterogeneous hospital systems. This proof-of-concept study aimed to evaluate the accuracy and query-level traceability of a commercial natural-language (NL) data-query platform implemented at a university hospital.

**Methods:** An on-premises data integration platform with an NL query interface (AIRE; Briya, Tel Aviv, Israel) was connected to the institutional health data platform across three tertiary-care hospitals under an approved pseudonymization and governance framework. Implementation comprised regulatory and governance approval, and IT deployment. Twenty-five NL prompts addressing cohort identification and characterization in an adult ICD-10 C71-coded brain-tumor cohort (n = 1,594) were evaluated against manually written reference SQL and checked for a corresponding executed query in the exported log. Responses were classified into five categories. Prompts with unanswered or incorrect responses were re-posed in a new thread.

**Results:** The platform was implemented over two years. Fifteen of 25 responses were exact (n = 11), concordant after definition alignment (n = 1), or incorrect but traceable from the logged SQL (n = 3); six were plausible but incorrect without a logged query, and four remained unanswered. No incorrect unlogged result recurred in a new thread. Across the initial and repeat evaluations, 22 of 23 logged responses were correct or traceable, whereas all eight unlogged responses were incorrect.

**Conclusions:** Implementation was feasible but required substantial local institutional preparation. The NL interface reproduced SQL-derived results when definitions were explicit, while query-level provenance and verification of the displayed result were critical for identifying unreliable outputs.

## Introduction

Retrospective clinical research depends on the ability to translate routinely collected clinical data inside a hospital into reproducible research cohorts. Although university hospitals increasingly provide centralized clinical data infrastructures [1,2], cohort identification often still requires project-specific database queries and support from informatics specialists [3]. Graphical feasibility tools let researchers check cohort availability without writing queries, but their usability remains limited [4], and the actual cohort still has to be built with database queries. Natural-language (NL) interfaces that translate clinical questions into executable database queries could therefore lower an important technical barrier to institutional research data and enable iterative on-demand querying without generating a new static data extract for each research question [5–8].

Large language model (LLM)–based text-to-SQL systems have increasingly been investigated for this purpose. Previous work has evaluated the translation of NL questions into structured database queries [5,6,9–11], while studies using hospital-derived tabular data suggest that LLM-based systems perform more reliably when they generate executable code than when they derive answers directly from the underlying data [12]. Tool-connected LLM systems querying clinical data warehouses have similarly shown high accuracy for comparatively simple retrieval tasks, whereas temporal and more complex operations remain challenging [13]. However, most evaluations have been performed using research databases, benchmark datasets, or experimental systems [5,6,9–13] rather than commercially deployed platforms embedded within routine hospital infrastructure. In such settings, performance must be assessed not only by whether a numerical result is correct, but also by whether its derivation can be independently verified [14–16].

For clinical research, this distinction is especially important. LLM-generated responses can be fluent but incorrect [17,18], vary between repeated interactions [19–21], and appear grounded without verifiable provenance [14]. Accordingly, evaluation of conversational data-query systems should consider both answer accuracy and query-level traceability. Whether a deployed platform meets both requirements in routine hospital infrastructure has not been well investigated so far.

Following calls for deployment-centered evaluation of clinical LLM systems [22], we implemented a commercially available NL clinical data-query platform within the institutional health data infrastructure of a German university hospital comprising three tertiary-care hospitals. We report the technical and organizational implementation process and evaluate the system in a neuro-oncological proof-of-concept study using manually written SQL as the reference. The primary aim of this study was to assess the platform’s query performance, defined as answer accuracy against the reference SQL and query-level traceability, in a real-world hospital deployment. Secondary analysis examined sources of discrepancy, first-prompt performance, and the reproducibility of failed or incorrect responses when queries were repeated in a new conversation thread without access to the previous conversational context.

## Methods

### Study design

We conducted an institutional implementation and subsequent single-center proof-of-concept evaluation of a commercially available NL data-query platform at Charité – Universitätsmedizin Berlin, a university medical center with three tertiary-care hospitals that share one clinical IT infrastructure and one institutional data platform, from October 2024 to September 2026. The study protocol was approved by the local institutional ethics committee (EA1/239/24).

### Institutional setting and data source

Relevant structured clinical data were accessed through the institutional Health Data Platform (HDP), a pseudonymized mirror of the clinical systems containing structured data from all three hospitals, including patient master data, encounters, diagnoses, and procedures. Data are queried with SQL through the Impala query engine. Access is governed by GDPR-compliant pseudonymization, role-based access control, project-specific approval, and audit mechanisms. The HDP holds relational tables rather than FHIR resources; the FHIR representation used by the platform was created during platform deployment (see below). Data integration centers of this kind were established at German university hospitals by the Medical Informatics Initiative, whose core data set is modeled in HL7 Fast Healthcare Interoperability Resources (FHIR) [1,2].

### Commercial platform implementation and architecture

The commercial data query platform (AIRE, described by the vendor as a scientific AI research environment; Briya, Tel Aviv, Israel) was deployed on a virtual server within the hospital network and connected directly to the institutional HDP. Implementation proceeded in two stages. The first stage comprised regulatory, data-protection, and governance approval. The second stage comprised installation and technical configuration on hospital infrastructure, and its FHIR layer was populated from the HDP tables without an export step. Vendor staff performed the installation and the data mapping. The language model and query components operated on-premises server, ensuring that no patient-level data left the hospital network. The platform was not modified during the study period.

### Platform operation and user interaction

Users interacted with the platform through an NL chat interface (Figure 1). The platform generated and executed SQL through the Trino query engine and requested clarification when a query was ambiguous. Each response was returned together with a short methodological note and the generated query. An exportable query log recorded the timestamp, SQL, row count, and execution status of each query (Supplemental Figure S1). This log served as the provenance channel of the study. A response without a generated query was identifiable on screen at the time of the answer and was confirmed in the exported log. The platform was evaluated without additional vendor-side instrumentation.

**Figure 1.**
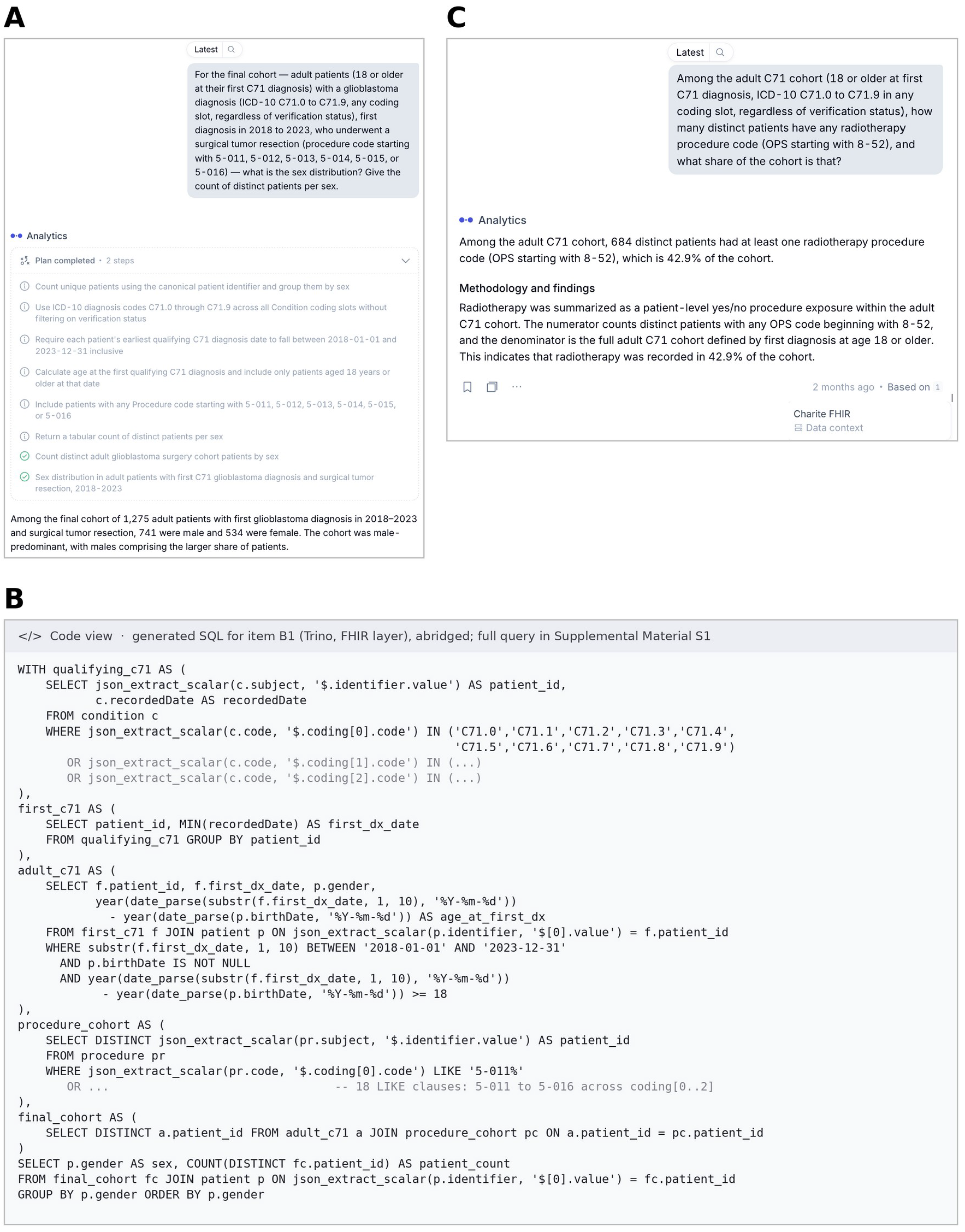
Query interface and provenance of the evaluated platform. (A) Logged item (B1, sex distribution of the resected cohort): the user prompt, the platform’s plan, and the narrative answer. (B) The generated SQL for this item (Trino, FHIR layer) as shown in the platform’s code view, abridged; the full query is in Supplemental Material S1. (C) Unlogged item (V13, radiotherapy exposure): the platform returned a numeric answer with a methodological explanation, but no plan or query was shown, and the answer was attributed only to the data context; the query log contains no data query for this prompt (reference 116 of 1,594 patients, 7.3%, versus 684, 42.9%, reported). The query activity log is shown in Supplemental Figure S1.

### Proof-of-concept evaluation

The proof-of-concept was designed to reflect a typical retrospective neuro-oncology cohort-selection workflow. The primary objective was to identify an adult cohort for a glioblastoma-oriented neuro-oncology use case, diagnosed between 2018 and 2023, and to further identify patients who underwent subsequent tumor resection.

A base cohort was defined as adults (18 years or older at first diagnosis) with any ICD-10 code from C71.0 to C71.9 in any coding slot, using C71 as a pragmatic proxy for glioblastoma. Because C71 is not specific to this tumor entity, the resulting cohort may represent a broader malignant brain-tumor population [23].

Additional stepwise cohort-construction items reflected the remaining predefined eligibility criteria of diagnosis period and surgical treatment, including definition of a subgroup of patients who underwent tumor resection. The resulting adult C71-coded cohort and the subgroup of patients who underwent tumor resection served as the reference cohorts for all items. Beyond cohort construction, characterization and variable-level queries were selected to test increasingly complex operations relevant to clinical cohort description and subgroup analysis.

The battery comprised 25 predefined items, each posed as an NL prompt (Table 2). The items were organized in three blocks: stepwise cohort construction (A1–A4: C71 → adult → first diagnosis 2018–2023 → resection), cohort characterization (B1–B4), and 17 additional variables of increasing complexity (V1–V17), ranging from grouped distributions to patient-level thresholds, event sequences, and subgroup statistics. Blocks B and V were evaluated in the full adult cohort, except B1, which used the resected sub-cohort.

The battery was run between July and September 2026 in a single chat thread, such that all preceding turns remained available in the platform’s conversational context. Eight prompts were posed in a second, parallel thread: two for the base count (A1), five attempts at the mean age (B2), and one aborted attempt (V12). Exploratory prompts preceded the battery and were not scored. When the platform asked a clarifying question or aborted, the prompt was refined by specifying operational definitions (fields, prefix matching, explicit date range), never by providing information about the expected result. The number of prompts required for each item was recorded. The response after final refinement was defined as the primary evaluation result.

To assess first-prompt performance separately, the response to the first prompt of each item was recorded as a correct number, an incorrect number, a clarifying question, or an abort. Numerical responses returned during intermediate steps without a corresponding logged query were recorded as unlogged responses.

### Reference queries and outcome definitions

For each predefined query item, the platform-generated result was compared with the result of a manually written reference SQL query. Because both workflows ultimately accessed data derived from the same institutional HDP source tables, manually written HDP SQL was used as the reference. Reference queries were written in Impala SQL by DD before the corresponding prompt was posed, so that their construction was not influenced by the platform-generated query or result. They represent a pre-specified operationalization of each item rather than an independent ground truth; their internal consistency was checked across the stepwise cohort items, and discrepancies were investigated as described below.

Discrepancies were assessed at the levels of query logic, patient-level cohort composition, and query provenance. Platform-generated SQL was reproduced in Impala to evaluate query logic; patient-level identifier exports were compared when cohort composition differed; and query-log timestamps were matched to the chat transcript to establish provenance. Medians were calculated outside the platform from patient-level exports. Response latency was defined as the interval between submission of a prompt and receipt of the platform’s response, based on the chat export. Each item was placed in one of five mutually exclusive categories (Table 1).

**Table 1.** Outcome categories for platform responses.

| No. | Category | Definition | Logged query |
|---|---|---|---|
| 1 | Exact | All values identical to the reference. | yes |
| 2 | Concordant after definition alignment | Deviation fully explained by a legitimate definitional choice visible in the SQL, such as the coding slot, the age computation, or prefix versus leaf codes. | yes |
| 3 | Incorrect but traceable | Deviation caused by a specific SQL clause that was not requested. | yes |
| 4 | Incorrect and unlogged | Value deviates and no executed query exists for it. | no |
| 5 | Not answered | Abort, explained abstention, or unresolved clarifying question. | — |

Categories are mutually exclusive and were assigned by two raters independently (Methods). Logged query = an executed data query for the response was present in the exported query log; category 4 is defined by its absence. Categories 1–3 are correct or traceable from the logged SQL; category 5 = no answer was generated.

A response was considered logged when the exported query log contained an executed data query between the prompt and response timestamps whose logic corresponded to the requested operation. Schema-information queries were excluded. Query timestamps were matched to the chat transcript to assign executed queries to individual responses.

The classification comprised 37 responses: 25 primary-thread items and 12 re-test items. Two investigators (DD and GG), working independently and blinded to each other’s classifications, assigned each response to one of the categories in Table 1. DD had run the battery, whereas GG had not been involved in the evaluation; GG received the prompts, platform answers, reference results, and logged SQL. Disagreements were resolved by discussion, and the consensus categories were used for all results; the independent ratings served only to quantify agreement. No hypothesis was prespecified. The analysis was descriptive, with results reported as counts within each category. Interrater agreement was assessed using percentage agreement and Cohen’s kappa [24]. Item-level classifications, prompts, generated and reference SQL, and matched log entries are provided in Supplemental Material S1.

Because no reporting guideline specifically addresses evaluations of NL clinical data-query interfaces, TRIPOD-LLM was consulted [25]. Applicable elements were reported that apply to a user-side evaluation of a deployed commercial system, including data source, task, evaluation protocol, human oversight, and limitations. Elements on model development and training data were not accessible to end users.

### Exploratory re-test

Immediately after completion of the primary thread, all items that had remained unanswered or had produced incorrect result were re-posed in a new chat thread without prior conversational context. Each prompt was posed verbatim, with one attempt per item. If the platform requested clarification, the same operational definitions used during the primary evaluation were provided. Apart from the absence of prior conversational context, the prompting procedure and operational definitions were kept unchanged.

## Results

### Implementation

The project was initiated in October 2024. Implementation required nearly two years, with regulatory approval and institutional IT deployment each accounting for roughly half of this period. Regulatory, data-protection, and governance approval took approximately nine months and included a data protection impact assessment and a two-stage information-security review, an initial risk mapping followed by a tailored risk assessment. Installation and technical configuration on hospital infrastructure started in July 2025 and was driven primarily by institutional prerequisites: virtual-server provisioning, VPN and network access, firewall clearance, and interoperability with legacy systems. Once server access was established, vendor-led installation, FHIR data mapping, and initial testing were completed within approximately two weeks. The first successful query was executed in June 2026, followed by about one month of institutional testing before the evaluation.

### Cohort characteristics

The adult C71 cohort comprised 1,594 patients with a mean age of 55.1 years at first diagnosis; 1,279 (80.2%) underwent tumor resection and 258 had a biopsy only. Among the 1,275 patients resected after a first diagnosis in 2018–2023, 741 were male and 534 female. Resection was coded on the day of the coded diagnosis in 49.5%, before it in 26.1%, and after it in 24.4% of patients. In-house death was documented for 101 patients (6.3%). All values are reference-SQL results (Table 2).

**Table 2.** Results of the 25-item battery in the primary thread and exploratory new-thread re-test.

| ID | Query | Prompts | Primary thread results / reference SQL | Logged | New thread results (12 re-posed items, 13 prompts) |
|---|---|---|---|---|---|
| A1 | C71 cohort | 2 | 1,880 / 1,880 | yes | — |
| A2 | + age ≥18 | 2 | 1,594 / 1,584<sup>1</sup> | yes | — |
| A3 | + first dx 2018–2023 | 7 | 1,587 / 1,587<sup>2</sup> | yes | — |
| A4 | + resection | 4 | 1,248 / 1,275<sup>3</sup> | yes | — |
| B1 | Sex distribution (resected) | 1 | 741/534 exact | yes | — |
| B2 | Mean age | 6 | 64.9 → 55.1 / 55.1 y | yes<sup>5</sup> | 53.2 y, n = 113<sup>6</sup>, logged |
| B3 | Mortality | 1 | 101 / 101 | yes | — |
| B4 | Top-15 procedures | 1 | 15/15 | yes | — |
| V1 | C71 subcodes | 4 | 10/10 | yes | — |
| V2 | Resection; biopsy only | 1 | 1,275; 117 / 1,279; 258<sup>4</sup> | no | explained abstention (decomposition offered) |
| V3 | Days dx→resection | 1 | 49.5/26.1/24.4% exact | yes | — |
| V4 | ≥2 resections | 1 | 34/95 / 220/1,279 | yes<sup>6</sup> | 13/83<sup>7</sup>, logged |
| V5 | First dx per year | 1 | Σ1,594 / Σ1,587 | no | aborted |
| V6 | Top-10 other diagnoses | 1 | 9/10 codes | yes | — |
| V7 | Age decades | 5 | failed; control 494 / 337 | no | 7/7 exact, logged |
| V8 | Sex × age band | 3 | 113 → 1,594 / 1,594 | yes | — |
| V9 | Cases per patient | 1 | median 1 / 1 | yes | — |
| V10 | Dx codes per patient | 2 | abstained / median 12 | — | median 13, concordant<sup>8</sup>, logged |
| V11 | MRIs per patient | 1 | aborted / median 2 | — | aborted |
| V12 | Chemotherapy 8-54 | 2 | 512 / 142 | no | aborted |
| V13 | Radiotherapy 8-52 | 2 | 684 / 116 | no | 116 exact, logged |
| V14 | Shunt 5-023 | 2 | 88 / 76 | no | 76 exact, logged |
| V15 | Days dx→death | 1 | median 231 / 43 | no | 93 d, n = 84<sup>9</sup>, logged |
| V16 | Deceased: sex, mean age | 1 | 35/66; 59.5/62.2 y exact | yes | — |
| V17 | Biopsy before resection | 2 | aborted / 23/56 | — | 23/56 exact, logged |

Prompts = user prompts per item in the thread of the scored answer, including answers to clarifying questions and verbatim repeats (B2: five further attempts in the parallel thread not counted; V12–V14 asked twice with identical results; new thread: 13 prompts for 12 items). <sup>1</sup> Concordant after definition alignment: all coding slots and calendar-year age; the reference rebuilt with these definitions also gives 1,594. <sup>2</sup> After an explicit date-range prompt; a naive prompt returned 1,594. <sup>3</sup> 48-code list vs prefix matching on 5-011 to 5-016; the 27 missing patients carried five-digit sub-codes or .X/.Y suffixes. 1,275 refers to the 2018–2023 cohort, 1,279 (V items) to the full adult cohort. <sup>4</sup> Both values unlogged. <sup>5</sup> First answer (64.9 y) unlogged, final answer logged. <sup>6</sup> Default verification-status filter in the logged query. <sup>7</sup> Default filter plus a self-added date restriction, disclosed in prose. <sup>8</sup> All three coding slots counted; the reference rebuilt accordingly gives 13. <sup>9</sup> Displayed for 84 patients, logged query returned 101 (display ≠ query). Categories are the consensus of two raters (Methods). Prompts, generated SQL, and reference SQL for every item are in Supplemental Material S1.

### Comparability of data access paths

The reference cohort comprised 1,594 adults with a C71 code, including 1,279 patients with tumor resection. When equivalent operational definitions were applied, the cohort-construction and characterization queries (A1–B4) yielded concordant patient counts across the platform and the reference SQL, with one exception. Two timing and subgroup items reproduced percentages and mean ages to the decimal. For the tumor-resection step (A4), a patient-level identifier comparison showed that the platform-derived cohort was a strict subset of the reference cohort; the discrepancy was attributable to code depth, with deeper OPS subcodes not captured by the platform’s explicit code list. No other discrepancy attributable to the data mapping was identified. Unit handling and the treatment of missing values were not evaluated beyond these concordances.

The exported query log comprised 100 entries: 30 schema lookups and 70 data queries. All 70 data queries could be assigned to a chat response or to a separate patient-identifier export, no response had a candidate query outside its prompt-to-response window, and chat and log timestamps agreed to the minute. When a single turn generated several queries (three items), their temporal sequence allowed unambiguous assignment. The log was therefore considered complete for the evaluated battery.

The two raters agreed on 30 of 37 responses (81%; Cohen’s κ = 0.74): 21 of 25 primary-thread responses (κ = 0.77) and 9 of 12 re-test responses (κ = 0.65). No disagreement crossed the boundary between categories 1–3 and 4–5. All seven disagreements concerned the distinction between a definitional choice and an unrequested restriction, or between an abort and an unlogged answer, and were resolved by discussion. Consensus categories are reported below and in Table 2.

Both access paths reflected the same characteristics of the routine data: a mixed C71 population (mean age 55 years, 22% under 40), in-house-only mortality of 6.3%, under-coded external radiotherapy and chemotherapy, and resection preceding the coded diagnosis in 26% of patients.

### Primary evaluation and failure modes

Of the 25 primary-thread items, 11 responses were exact, one was concordant after definition alignment, three were incorrect but traceable, six were incorrect and unlogged, and four were not answered (Table 2; Figure 2a). Exact matches, including the generated SQL, covered counts and rankings (mortality, the top-15 procedure codes, the sex and C71-subcode distributions) as well as more complex operations: a sex × age cross-tabulation, a per-patient encounter summary, a subgroup analysis of deceased patients, and a diagnosis-to-resection timing distribution reproduced to the decimal (49.5% same day, 26.1% before, and 24.4% after the coded diagnosis; n = 1,279). Stepwise cohort construction returned 1,880, 1,594, 1,587, and 1,248 patients (reference 1,880, 1,584, 1,587, and 1,275; the reference resection count refers to first diagnoses in 2018–2023, whereas the 1,279 used for the V items is the resection subgroup of the full adult cohort). The concordant item was the adult cohort (A2): the platform counted C71 in all coding slots and computed age by calendar year, whereas the reference used the exact date of birth, and the difference of 10 patients was visible in the SQL. The three traceable deviations each arose from a specific clause in the logged SQL: a list of 48 leaf codes instead of prefix matching on 5-011 to 5-016 (A4), a NULL code ranked first in a top-10 diagnosis ranking so that 9 of 10 codes matched (V6), and a default verification-status filter in the re-resection rate (V4).

**Figure 2.**
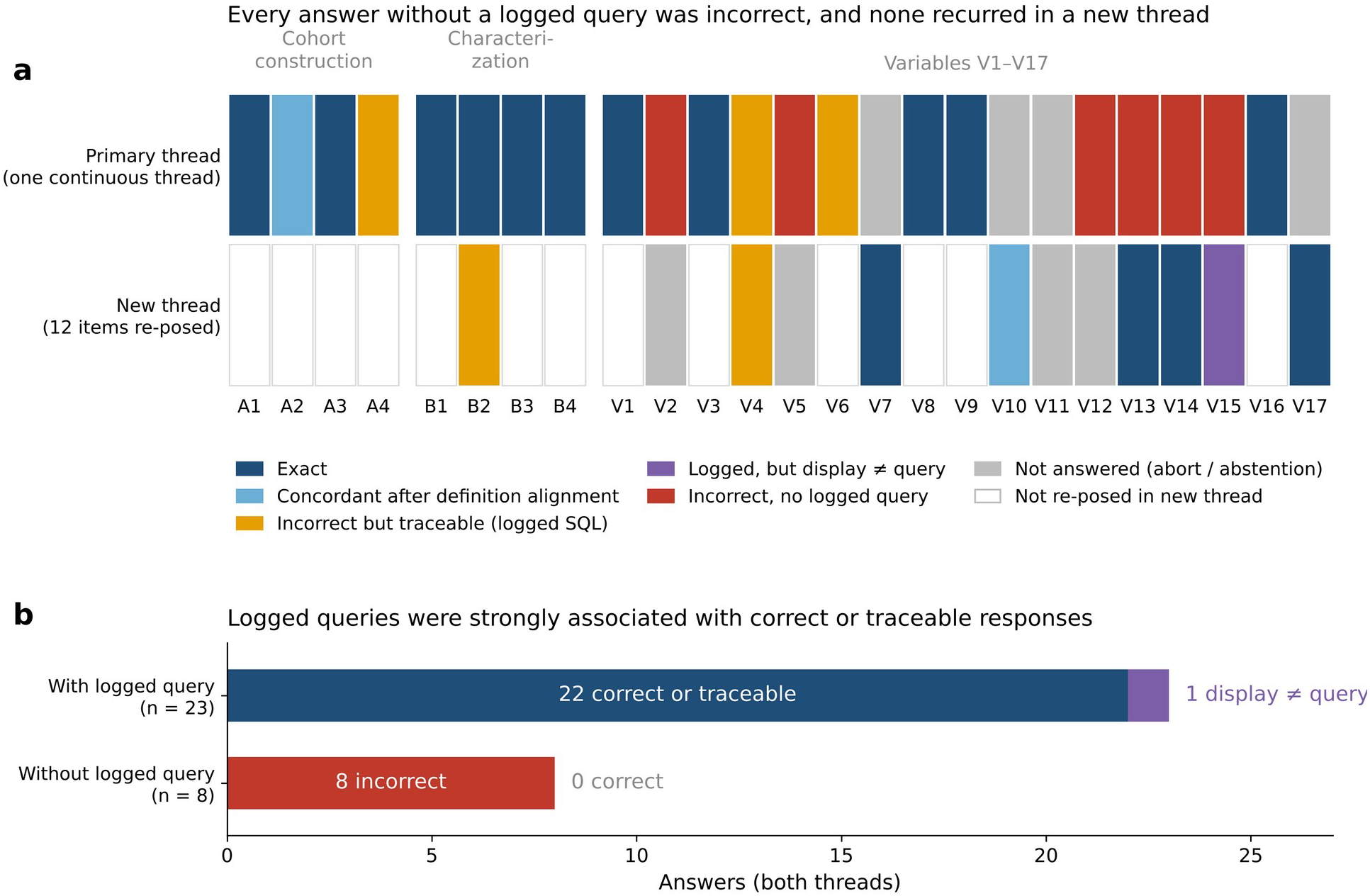
Provenance and outcome across the 25-item battery. (a) Outcome of every item in the primary thread (one continuous thread over six weeks) and, for the 12 items re-posed verbatim, in a new thread without prior context; white cells = not re-posed. (b) Answers pooled across both threads by whether the exported query log contained an executed query for the answer: 22 of 23 logged answers were correct or traceable from their SQL (the exception being the display–query mismatch in V15); all eight numbers returned without a logged query — six final answers and two intermediate values (64.9 years; 494) — were incorrect. Categories as defined in Methods; item-level detail in Table 2 and Supplemental Material S1.

First-prompt performance was lower than the final result. Six of the 25 items were answered exactly by the first prompt (B1, B3, B4, V3, V9, V16) and two more with a traceable deviation (A4, V6); three began with a clarifying question, seven with an abort or a response without a number, and seven with an incorrect number that was never corrected in the primary thread (V2, V4, V5, V12–V15). Of the 12 items finally scored exact or concordant, nine were correct at their first numeric answer; in three, a refined prompt corrected the first number (1,594 → 1,587 once the date range was spelled out; 64.9 → 55.1 years after a summary-style prompt; 113 → 1,594 after “do not filter on verification status”). First-prompt accuracy was thus 6 of 25 by the strictest criterion and 9 of 25 at the first number returned, compared with 12 of 25 after refinement. Every refinement specified a criterion, never a value, and items in categories 1–3 took a median of one prompt (range 1–7).

The platform asked a clarifying question in seven items, offered alternative code-set definitions for resection, and ended four items with an abort or an explained abstention that proposed alternative variables. Every logged deviation could be traced to a specific clause. Two errors, the re-resection rate (V4) and, at an intermediate step, the sex × age cross-tabulation (V8), shared one cause: a default verification-status filter (verificationStatus = ‘G’) that restricts cohorts to confirmed diagnoses and, because Condition.verificationStatus is an optional modifier element in FHIR R4, also drops conditions in which the element is absent [26]. The filter was applied even when the prompt said “regardless of verification status”; an explicit “do not filter on verification status” clause removed it reliably thereafter.

Six items were answered with fluent methodological prose but without any query, in the log or on screen, and all six deviated from the reference (Table 2), for example biopsy-only patients 117 (reference 258), chemotherapy 512 patients, 32.1% (142, 8.9%), and a median diagnosis-to-death interval of 231 days (43 days for in-house documented deaths). The six yearly first-diagnosis values were identical, year for year, to a per-year breakdown logged earlier in the thread, and the three code-family counts (V12–V14) returned the same unlogged numbers when requested again days later. Two further unlogged numbers appeared at intermediate steps (B2, V7), one of which exactly matched a value previously reported for a differently defined age band. When asked for the SQL behind an unlogged answer, the platform stated that no query was included in the results it had received. The parallel thread provided the counter-example: a mean age of 52.3 years was incorrect but logged, and its SQL showed that the query had run on all adult patients without the C71 restriction.

### Exploratory re-test and query provenance

In the new thread, none of the incorrect numbers recurred (Table 2; Figure 2a). Four items were exact and logged at the first attempt: two code-family counts, the age-decade distribution, and the biopsy-before-resection sequence. The per-patient count of diagnosis codes was concordant after definition alignment (median 13 vs 12): the platform counted distinct codes across all three coding slots, whereas the reference counted the primary ICD code only, and re-running the reference over all three slots reproduced the platform result exactly. Two items were incorrect but traceable, the mean age (53.2 years, n = 113) and the re-resection rate (13/83), both carrying the default filter; the latter additionally carried a disclosed restriction to procedures after the diagnosis date. One item (V15) was logged but did not match its own query: the diagnosis-to-death query returned 101 distinct patients, the in-house mortality count, yet statistics were displayed for 84 patients, a reduction with no counterpart in the query or the log. Four items were not answered, one explained abstention and three aborts. No answer lacked a logged query.

Across both threads, 8 of the 37 evaluated responses were aborts or abstentions rather than numbers, and every unlogged number occurred in the primary thread. Two structurally identical code-family prompts diverged in the new thread (radiotherapy exact, chemotherapy aborted), and the mean-age question yielded four values across threads: 64.9 unlogged, and 55.1, 53.2, and 52.3 logged, the last two traceable from their SQL. In sum, 22 of 23 logged answers were correct or traceable, the exception being the display–query mismatch, whereas all six unlogged answers and both intermediate unlogged numbers were incorrect (Figure 2b).

### Response latency

Numeric responses with a logged query had substantially longer response times than unlogged numeric responses. The 27 numeric responses with a logged query arrived after a median of 64 s (interquartile range 57–81 s; range 45–371 s), with one exception at 3 s, the incorrect mean age of 52.3 years in the parallel thread. All 15 numeric answers without a logged query arrived within 3–6 s, as did all 25 aborts and all 8 clarifying questions. With that single exception, every number that appeared within seconds had no executed query behind it.

## Discussion

In this real-world proof-of-concept study, we evaluated the institutional implementation, accuracy, and query-level traceability of a commercial natural-language data-query platform connected to a large hospital data platform. The platform enabled NL querying of institutional clinical data, but response reliability was strongly associated with the presence of an executed, traceable database query. When a corresponding SQL query was logged, results were almost always correct, or deviations could be explained from the executed query; numerical responses without a logged query were consistently incorrect. Prompt wording and operational definitions also influenced performance, and several errors did not recur when the same items were re-posed in a new chat thread. Implementation was feasible but required nearly two years, with most of the effort attributable to regulatory, governance, and institutional IT requirements rather than software installation.

A key implementation finding was that integration through a single institutional data platform provided a practical entry point for deployment across a multi-site university medical center under the GDPR. Rather than connecting the NL interface separately to multiple clinical source systems, the platform could build on the existing HDP and its established pseudonymization, access-control, approval, and audit mechanisms. That governance framework is what makes it possible to offer an NL interface to clinician-researchers at all, and it reduced the technical integration burden once the institutional infrastructure was in place. The benefit arose from coupling the NL interface with a standardized FHIR layer built on this centralized platform; wider adoption of FHIR in hospital data infrastructures would reduce the integration effort for such systems. Existing secondary-use data platforms may therefore provide an effective foundation for introducing NL data-query tools in regulated hospital environments. Extension to further departments and data sources is part of the ongoing implementation and lies outside this report.

Once the NL interface was available, the question was what a clinician can rely on. In a fully on-premises, regulated deployment, the platform reproduced SQL-derived cohort results for most operation types when definitions were explicit and the thread was fresh. Performance was not determined by the type of operation requested: the platform reproduced reference results for basic descriptive tasks and for cross-tabulations, per-patient summaries, subgroup analyses, and event-sequence queries, whereas several failures occurred for comparatively simple counts. Both the NL interface and the reference SQL also reflected the same limitations of routine clinical data. The observed in-house mortality of 6.3% is implausibly low for a disease with a median survival of about 14.6 months under standard chemoradiation [27], indicating incomplete capture of outcomes outside the institution. An NL interface surfaces such data-quality limitations as readily as conventional SQL, but it does not necessarily identify or contextualize them for the user.

The importance of query provenance is consistent with text-to-SQL approaches in which the model generates or invokes an executable query rather than producing an answer directly [6,12,13]. Recent systems expose the executed SQL alongside the result [7], and agentic approaches add schema grounding and iterative query correction [8,28]. Our findings concern the deployment-side counterpart: the executed query must remain visible and auditable to the user. The platform’s clarification behavior served the same purpose. It requested definitions, offered alternative code-set interpretations, and in some instances disclosed restrictions it had introduced itself. Because NL questions may permit several valid SQL interpretations [29], such clarification makes otherwise implicit analytical choices explicit.

The second central finding was the association between traceability and thread state. Every numerical response without a logged query was incorrect, whereas re-posing the same items in a new thread yielded logged correct responses, traceable errors, or explicit abstentions, but no unlogged number. Several observations suggest that accumulated conversation context led the platform to reuse earlier values instead of executing a new query, and some of these unlogged outputs were clinically plausible and would have been difficult to question without the reference SQL. This reading agrees with the literature. Sequential clinical text-to-SQL accuracy depends on how conversation history is represented [10], which is consistent with the same items succeeding once the history was removed. Frontier LLMs lose reliability when task information accumulates across turns and commit to early assumptions [21], which matches the reuse of a logged per-year breakdown as the unlogged answer to a later question. Multi-turn text-to-SQL models gain accuracy when schema elements and prior SQL are tracked explicitly rather than left in an ever-growing context [30]; our six-week thread is an extreme version of the untracked setting. Non-deterministic execution, which we observed directly, cannot be ruled out for individual items; it is the pattern across all 12 re-posed items that carries the argument. The finding extends the provenance gap described for citations [14] to structured analytics, and it means that failures that at first appeared to be intrinsic limits of the platform were artifacts of the thread.

Three further behaviors matter for practice. First, a default verification-status filter restricting cohorts to confirmed diagnoses was applied unless the prompt said “do not filter”; “regardless of verification status” did not prevent it in three instances. Such a default may suit clinical use, but in research it should be explicit and overridable, because defaults embedded in generated SQL can be overlooked even when the query is displayed. Second, structurally identical prompts were executed non-deterministically, in line with the repeated-run variability reported for LLMs in other clinical tasks [20]. Third, observed once, the presentation layer deduplicated rows and then lost some, so a logged query is necessary but not sufficient. Reproducibility of routine-data studies depends on a complete specification of the cohort-extraction logic [15], and provenance capture has long been proposed as a requirement of learning health systems [16]. Provenance does not guarantee correctness, but it turns opaque errors into traceable ones: the default filter was identified from one SQL line and fixed with one clause, whereas the unlogged chemotherapy share remained unexplained.

Several practical measures follow. Users should keep threads short and separated by topic, state every definition in every prompt, and accept no number without a displayed query; its absence, together with a response within seconds, was the most reliable warning sign we found. They should keep the executed query for any number they intend to use, check that the displayed count follows from it, and repeat critical queries in a new thread. Platform developers should flag the absence of an executed query as a reliability signal and abstain instead of answering, in the spirit of reliability-scored text-to-SQL [9] and calibrated abstention [31], and reconcile presentation-layer counts with query row counts. Criteria that exist only in free text are poorly served by structured-only retrieval; precision among the top five retrieved patients rose from 0.54 to 0.90 in one proof of concept when clinical text processed with natural language processing (NLP) was added [32]. We plan to evaluate free-text retrieval once report text becomes available on the platform, with the same provenance requirement for every source added.

The study has limitations. It covered a single site, vendor, cohort, and platform version. The reference standard was query execution, not clinical truth, and the mapping was validated by comparing outputs rather than field by field; where a prompt left a definition open, the deviation could lie on the reference side, as in the per-patient code count, where the platform’s reading was the more complete one. Patient-level cohort composition was compared only for the resection step (A4), so compositional differences that leave a count unchanged would not have been detected. The battery was descriptive, developed in three blocks over six weeks, and not pre-specified. Two raters classified the items independently, but neither was blinded to the reference values, and the first rater had run the battery; all classifications can be audited from Supplemental Material S1. Each item had one phrasing, so sensitivity to paraphrase is unknown, and the re-test was post hoc with a single attempt per item. Future studies should prospectively assess repeated-run reproducibility, robustness to prompt wording, and the effect of conversational context under preregistered protocols with blinded evaluation.

## Conclusion

Integration of a natural-language data-query platform through an institutional health data platform was feasible in a German university hospital, although implementation required substantial regulatory, governance, and IT preparation. The NL interface reproduced SQL-derived cohort results across a broad range of query types, but performance was sensitive to operational definitions and conversational context. Query-level provenance emerged as a key requirement for reliable use: logged queries made numerical outputs auditable and allowed discrepancies to be traced to specific query logic, whereas unlogged numerical responses were consistently incorrect. However, the presence of a logged query alone did not guarantee agreement between query output and the displayed result. Reliable clinical-research use therefore requires both transparent query provenance and verification that displayed results accurately reflect the underlying executed query.

## Statements and declarations

## Data Availability

Prompts, generated and reference SQL, and the matched query-log entries for every item are provided as Supplemental Material S1. The full chat transcripts and the query-log export are available from the corresponding author on reasonable request. Patient-level data cannot be shared.

## Acknowledgements

we thank Talia Friedman and Chagai Yariv (Briya) for the data mapping and the installation of the platform, and Thilo Wetzer, Florian Seidel, and Camille Guinemer (Berlin Institute of Health at Charité) for setting up the platform server, connecting it to the HDP, and providing the HDP data.

## Ethical considerations

the study was approved by the Ethics Committee of Charité – Universitätsmedizin Berlin (EA1/239/24; approval of 29 October 2024, conditions confirmed fulfilled on 7 January 2025) as a retrospective analysis of pseudonymized routine data without individual informed consent; no patient-level data left the hospital network.

## Data availability

prompts, generated and reference SQL and the matched query-log entries for every item are provided as Supplemental Material S1; the full chat transcripts and query-log export are available from the corresponding author on reasonable request; patient-level data cannot be shared.

## Author contributions (CRediT)

DD — conceptualization, methodology, investigation, data curation, formal analysis, visualization, writing – original draft. PB — project administration (platform implementation), resources, software, investigation, writing – review and editing. AD — methodology, writing – review and editing. GG — formal analysis (independent second rating), writing – review and editing. MPW — supervision, writing – review and editing. JN — conceptualization, supervision, project administration, writing – review and editing.

## AI use

language-model assistance (Claude, Anthropic) was used for drafting, editing and reference checking under author supervision; all content was verified by the authors.

## References

1. Semler SC, Wissing F, Heyder R. German Medical Informatics Initiative. Methods Inf Med 2018; 57(S 01): e50–e56. doi:10.3414/ME18-03-0003.

2. Ammon D, Kurscheidt M, Buckow K, et al. [Interoperability working group: core dataset and information systems for data integration and data exchange in the Medical Informatics Initiative]. Bundesgesundheitsblatt Gesundheitsforschung Gesundheitsschutz 2024; 67(6): 656–667. German. doi:10.1007/s00103-024-03888-4.

3. Dobbins NJ, Spital CH, Black RA, et al. Leaf: an open-source, model-agnostic, data-driven web application for cohort discovery and translational biomedical research. J Am Med Inform Assoc 2020; 27(1): 109–118. doi:10.1093/jamia/ocz165.

4. Schüttler C, Prokosch HU, Sedlmayr M, Sedlmayr B. Evaluation of three feasibility tools for identifying patient data and biospecimen availability: comparative usability study. JMIR Med Inform 2021; 9(7): e25531. doi:10.2196/25531.

5. Wang P, Shi T, Reddy CK. Text-to-SQL generation for question answering on electronic medical records. In: Proceedings of The Web Conference 2020 (WWW ‘20). p. 350–361. doi:10.1145/3366423.3380120.

6. Lee G, Hwang H, Bae S, et al. EHRSQL: a practical text-to-SQL benchmark for electronic health records. In: NeurIPS 2022 Datasets and Benchmarks Track. arXiv:2301.07695.

7. Al Attrach R, Moreira P, Fani R, et al. M3: conversational LLMs simplify secure clinical data access, understanding, and analysis. PLOS Digit Health 2026; 5(9): e0001671. doi:10.1371/journal.pdig.0001671.

8. Kulkarni P, Kanhere A, Yi PH, Parekh VS. Text2Cohort: facilitating intuitive access to biomedical data with natural language cohort discovery. arXiv:2305.07637 (2023).

9. Lee G, Kweon S, Bae S, Choi E. Overview of the EHRSQL 2024 shared task on reliable text-to-SQL modeling on electronic health records. In: Proceedings of the 6th Clinical Natural Language Processing Workshop (ClinicalNLP), NAACL 2024. p. 644–654. doi:10.18653/v1/2024.clinicalnlp-1.62.

10. Ryu J, Cho S, Lee G, Choi E. EHR-SeqSQL: a sequential text-to-SQL dataset for interactively exploring electronic health records. In: Findings of the Association for Computational Linguistics: ACL 2024. p. 16388–16407. doi:10.18653/v1/2024.findings-acl.971.

11. Park J, Fang Y, Ta C, et al. Criteria2Query 3.0: leveraging generative large language models for clinical trial eligibility query generation. J Biomed Inform 2024; 154: 104649. doi:10.1016/j.jbi.2024.104649.

12. Klang E, Sorin V, Korfiatis P, et al. Large language models are poor clinical administrators: an evaluation of structured queries in real-world electronic health records. PLOS Digit Health 2026; 5(5): e0001326. doi:10.1371/journal.pdig.0001326.

13. Masayoshi K, Hashimoto M, Yokoyama R, et al. EHR-MCP: real-world evaluation of clinical information retrieval by large language models via Model Context Protocol. arXiv:2509.15957 (2025).

14. Ahmed MS, Dusanic M, Kirschner MN, et al. The provenance gap in clinical AI: evidence-traceable temporal knowledge graphs for rare disease reasoning. arXiv:2604.17114 (2026).

15. Wang SV, Sreedhara SK, Schneeweiss S, et al. Reproducibility of real-world evidence studies using clinical practice data to inform regulatory and coverage decisions. Nat Commun 2022; 13(1): 5126. doi:10.1038/s41467-022-32310-3.

16. Curcin V. Embedding data provenance into the Learning Health System to facilitate reproducible research. Learn Health Syst 2017; 1(2): e10019. doi:10.1002/lrh2.10019.

17. Zhou K, Hwang JD, Ren X, Sap M. Relying on the unreliable: the impact of language models’ reluctance to express uncertainty. In: Proceedings of the 62nd Annual Meeting of the Association for Computational Linguistics (Volume 1: Long Papers); 2024. p. 3623–3643. doi:10.18653/v1/2024.acl-long.198.

18. Asgari E, Montaña-Brown N, Dubois M, et al. A framework to assess clinical safety and hallucination rates of LLMs for medical text summarisation. NPJ Digit Med 2025; 8(1): 274. doi:10.1038/s41746-025-01670-7.

19. Ouyang S, Zhang JM, Harman M, Wang M. An empirical study of the non-determinism of ChatGPT in code generation. ACM Trans Softw Eng Methodol 2025; 34(2): 1–28. doi:10.1145/3697010.

20. Shyr C, Ren B, Hsu C-Y, et al. A statistical framework for evaluating the repeatability and reproducibility of large language models. medRxiv 2025 [preprint]; version 3 posted 25 March 2026. doi:10.1101/2025.08.06.25333170.

21. Laban P, Hayashi H, Zhou Y, Neville J. LLMs get lost in multi-turn conversation. In: Proceedings of the International Conference on Learning Representations (ICLR) 2026. arXiv:2505.06120.

22. Unell A, Fuentes M, Li B, et al. Deployment-centered evaluation: predicting query-level rejection risk in a clinical LLM system. arXiv:2606.12702 (2026).

23. Ostrom QT, Price M, Neff C, et al. CBTRUS statistical report: primary brain and other central nervous system tumors diagnosed in the United States in 2016–2020. Neuro Oncol 2023; 25(Suppl 2): iv1–iv99. doi:10.1093/neuonc/noad149.

24. Cohen J. A coefficient of agreement for nominal scales. Educ Psychol Meas 1960; 20(1): 37–46. doi:10.1177/001316446002000104.

25. Gallifant J, Afshar M, Ameen S, et al. The TRIPOD-LLM reporting guideline for studies using large language models. Nat Med 2025; 31(1): 60–69. doi:10.1038/s41591-024-03425-5.

26. HL7 International. FHIR Release 4 (v4.0.1): Resource Condition – Condition.verificationStatus, https://hl7.org/fhir/R4/condition.html (2019, accessed 4 September 2026).

27. Stupp R, Mason WP, van den Bent MJ, et al. Radiotherapy plus concomitant and adjuvant temozolomide for glioblastoma. N Engl J Med 2005; 352(10): 987–996. doi:10.1056/NEJMoa043330.

28. Lee K, Hong S, Park J, et al. EMR-AGENT: automating cohort and feature extraction from EMR databases. arXiv:2510.00549 (2025).

29. Bhaskar A, Tomar T, Sathe A, Sarawagi S. Benchmarking and improving text-to-SQL generation under ambiguity. In: Proceedings of the 2023 Conference on Empirical Methods in Natural Language Processing; 2023. p. 7053–7074. doi:10.18653/v1/2023.emnlp-main.436.

30. Chen B, Shi S, Luo Y, et al. Track-SQL: enhancing generative language models with dual-extractive modules for schema and context tracking in multi-turn text-to-SQL. In: Proceedings of the 2025 Conference of the Nations of the Americas Chapter of the Association for Computational Linguistics: Human Language Technologies (Volume 1: Long Papers); 2025. p. 10690–10708. doi:10.18653/v1/2025.naacl-long.536.

31. Chen K, Chen Y, Yu X, Koudas N. Reliable text-to-SQL with adaptive abstention. Proc ACM Manag Data (SIGMOD) 2025. doi:10.1145/3709719.

32. Liu S, Wang Y, Wen A, et al. Implementation of a cohort retrieval system for clinical data repositories using the Observational Medical Outcomes Partnership Common Data Model: proof-of-concept system validation. JMIR Med Inform 2020; 8(10): e17376. doi:10.2196/17376.

