## Supplemental Material S1 for "Natural-language cohort queries on a hospital data platform: accuracy and provenance in a real-world evaluation"

### Supplemental Material S1 (v1.3) — Prompts, generated SQL and reference SQL for all 25 items

Desser D, Brunecker P, Dell'Orco A, Galperin G, Wattjes MP, Nawabi J. Natural-language cohort queries on a hospital data platform: accuracy and provenance in a real-world evaluation (Brief Report, DIGITAL HEALTH). Compiled from the exported platform query log (100 entries, 4 July–1 September 2026), the test matrix and the chat transcripts. For each item: the NL prompt(s) as posed (clarifying exchanges in square brackets), the platform's displayed result versus the Impala reference, the log entry/entries matched to the answer (timestamp, status, row count) with the generated SQL verbatim, and the hand-written Impala reference SQL. 'No query logged' = no data query with a timestamp inside the prompt–answer window (schema lookups disregarded). Re-test entries (new thread, 1 September 2026) follow the primary entry where applicable. Each item is preceded by its outcome classification as reported in Table 2 of the main text, followed by the independent ratings of the two raters (DD, GG) and the consensus category (1 Exact, 2 Concordant after definition alignment, 3 Incorrect but traceable, 4 Incorrect and unlogged, 5 Not answered; categories defined in Table 1 of the main text). Version 1.2 (16 October 2026): reference SQL for A4, B1 and B4 replaced by the cohort-level queries actually used (see notes in the respective items), A2 strict reference added, second reference variant for V10 (re-test) added, ratings added. Version 1.3 (5 November 2026): table numbering updated after the addition of Table 1 (outcome categories) to the main text, the battery table is now Table 2; code glossary added; A1 reference SQL harmonized with A2–A4 (both diagnosis columns), result unchanged.

Code glossary. ICD-10-GM C71.0–C71.9, malignant neoplasm of the brain: C71.0 cerebrum except lobes and ventricles, C71.1 frontal lobe, C71.2 temporal lobe, C71.3 parietal lobe, C71.4 occipital lobe, C71.5 cerebral ventricle, C71.6 cerebellum, C71.7 brain stem, C71.8 overlapping lesion, C71.9 brain, unspecified. OPS (German procedure classification): 5-011 to 5-016 incision, excision and destruction of intracranial tissue (used as tumor resection; prefix match in the reference, leaf codes 5-011.0 to 5-016.7 in the platform’s list, see A4); 1-510 and 1-511 biopsy of brain tissue without incision; 5-023 cerebrospinal fluid shunt procedures; 8-52 radiotherapy; 8-54 cytostatic chemotherapy; 3-820 MRI of the skull. HDP columns: c_diagnose_1 and c_diagnose_2 are the first and second ICD-10 coding slots of a case diagnosis (coding[0] and coding[1] in the platform’s FHIR Condition resource, coding[2] = c_orphacode); c_gewissheit is the verification status (Condition.verificationStatus, ‘G’ = confirmed).

#### A1 — GBM diagnosis (ICD-10 C71.0-C71.9)

*Outcome (Table 2): primary thread — exact; query logged: yes. Ratings DD / GG / consensus: 1 / 1 / 1.*

**Prompt(s) as posed:**

How many patients have a glioblastoma diagnosis (ICD-10 C71.0 to C71.9 in any coding slot)? Include all diagnoses regardless of verification status. Return one total count of distinct patients.

*[Platform asked whether to count by patient ID only or identify patients from diagnosis codes first.]*

Identify patients with glioblastoma from the diagnosis codes first: any Condition with ICD-10 code C71.0 to C71.9 in any coding slot (coding[0], coding[1], or coding[2]), regardless of verification status. Then count the distinct patients. Return one total count.

Platform result: 1880 | Impala reference: 1880

*[v1.2 note: the scored answer (1880, logged) comes from the parallel thread (prompt 22:56, answer 22:58, log entry 22:59 on 31 July 2026). The same prompt had been posed in the primary thread at 22:50 and was answered with 1594 without a logged query; this unlogged intermediate number is counted among the eight unlogged numbers reported in Results.] [v1.3 note: the reference SQL now filters both diagnosis columns, consistent with A2–A4; the earlier version used c_diagnose_1 only. Re-run on 5 November 2026: n = 1,880, unchanged.]*

**Generated SQL, primary thread — log entry Jul 31 2026 22:59 · status: data available · rows: 1**

```
SELECT COUNT(DISTINCT patient_id) AS total_distinct_patients
FROM (
    SELECT json_extract_scalar(c.subject, '$.identifier.value') AS patient_id
    FROM condition c
    WHERE (
        json_extract_scalar(c.code, '$.coding[0].code') LIKE 'C71.0%'
        OR json_extract_scalar(c.code, '$.coding[0].code') LIKE 'C71.1%'
        OR json_extract_scalar(c.code, '$.coding[0].code') LIKE 'C71.2%'
        OR json_extract_scalar(c.code, '$.coding[0].code') LIKE 'C71.3%'
        OR json_extract_scalar(c.code, '$.coding[0].code') LIKE 'C71.4%'
        OR json_extract_scalar(c.code, '$.coding[0].code') LIKE 'C71.5%'
        OR json_extract_scalar(c.code, '$.coding[0].code') LIKE 'C71.6%'
        OR json_extract_scalar(c.code, '$.coding[0].code') LIKE 'C71.7%'
        OR json_extract_scalar(c.code, '$.coding[0].code') LIKE 'C71.8%'
        OR json_extract_scalar(c.code, '$.coding[0].code') LIKE 'C71.9%'
        OR json_extract_scalar(c.code, '$.coding[1].code') LIKE 'C71.0%'
```

```
        OR json_extract_scalar(c.code, '$.coding[1].code') LIKE 'C71.1%'
        OR json_extract_scalar(c.code, '$.coding[1].code') LIKE 'C71.2%'
        OR json_extract_scalar(c.code, '$.coding[1].code') LIKE 'C71.3%'
        OR json_extract_scalar(c.code, '$.coding[1].code') LIKE 'C71.4%'
        OR json_extract_scalar(c.code, '$.coding[1].code') LIKE 'C71.5%'
        OR json_extract_scalar(c.code, '$.coding[1].code') LIKE 'C71.6%'
        OR json_extract_scalar(c.code, '$.coding[1].code') LIKE 'C71.7%'
        OR json_extract_scalar(c.code, '$.coding[1].code') LIKE 'C71.8%'
        OR json_extract_scalar(c.code, '$.coding[1].code') LIKE 'C71.9%'
        OR json_extract_scalar(c.code, '$.coding[2].code') LIKE 'C71.0%'
        OR json_extract_scalar(c.code, '$.coding[2].code') LIKE 'C71.1%'
        OR json_extract_scalar(c.code, '$.coding[2].code') LIKE 'C71.2%'
        OR json_extract_scalar(c.code, '$.coding[2].code') LIKE 'C71.3%'
        OR json_extract_scalar(c.code, '$.coding[2].code') LIKE 'C71.4%'
        OR json_extract_scalar(c.code, '$.coding[2].code') LIKE 'C71.5%'
        OR json_extract_scalar(c.code, '$.coding[2].code') LIKE 'C71.6%'
        OR json_extract_scalar(c.code, '$.coding[2].code') LIKE 'C71.7%'
        OR json_extract_scalar(c.code, '$.coding[2].code') LIKE 'C71.8%'
        OR json_extract_scalar(c.code, '$.coding[2].code') LIKE 'C71.9%'
    )
) matched
```

**Reference SQL (Impala, HDP):**

```
SELECT count(DISTINCT c_patnr) AS n
FROM db_tumpseudoprog_prepared.it_ishmed_diagnose
WHERE c_diagnose_1 LIKE 'C71%' OR c_diagnose_2 LIKE 'C71%';
```

#### A2 — + age >= 18 at first diagnosis

*Outcome (Table 2): primary thread — concordant after definition alignment; query logged: yes. Ratings DD / GG / consensus: 2 / 1 / 2.*

**Prompt(s) as posed:**

How many patients with a glioblastoma diagnosis (ICD-10 C71.0 to C71.9) were at least 18 years old at the time of their first C71 diagnosis? Include all diagnoses regardless of verification status.

*[Platform asked which column to use for age at first diagnosis or whether to derive it.]*

Derive it from the birth date and the diagnosis date: use the patient's date of birth and the date of their earliest (first) C71 diagnosis, and compute the age at that first diagnosis.

Platform result: 1594 | Impala reference: 1594 with the platform's definitions (all coding slots, calendar-year age); strict reference with age from the exact date of birth: 1584

**Generated SQL, primary thread — log entry Jul 31 2026 11:31 · status: data available · rows: 1**

```
SELECT COUNT(DISTINCT q.patient_id) AS patient_count
FROM (
    SELECT
        first_c71.patient_id,
        first_c71.first_c71_recorded_date,
        p.birthDate
    FROM (
        SELECT
            c.patient_id,
            MIN(c.recorded_dt) AS first_c71_recorded_date
        FROM (
            SELECT
                json_extract_scalar(cond.subject, '$.identifier.value') AS patient_id,
                date_parse(substr(cond.recordedDate, 1, 19), '%Y-%m-%dT%H:%i:%s') AS recorded_dt
            FROM condition cond
            WHERE
                json_extract_scalar(cond.code, '$.coding[0].code') LIKE 'C71%'
                OR json_extract_scalar(cond.code, '$.coding[1].code') LIKE 'C71%'
                OR json_extract_scalar(cond.code, '$.coding[2].code') LIKE 'C71%'
        ) c
        WHERE c.recorded_dt IS NOT NULL
        GROUP BY c.patient_id
    ) first_c71
    JOIN patient p
      ON first_c71.patient_id = json_extract_scalar(p.identifier, '$[0].value')
    WHERE date_diff(
            'year',
            date_parse(substr(p.birthDate, 1, 10), '%Y-%m-%d'),
            CAST(first_c71.first_c71_recorded_date AS date)
          ) >= 18
) q
```

**Reference SQL (Impala, HDP):**

```
WITH c71 AS (
  SELECT c_patnr, min(c_diagnose_timestamp) AS first_dx
  FROM db_tumpseudoprog_prepared.it_ishmed_diagnose
  WHERE c_diagnose_1 LIKE 'C71%' OR c_diagnose_2 LIKE 'C71%'
  GROUP BY c_patnr),
bd AS (SELECT DISTINCT c_patnr, c_birthdate FROM db_tumpseudoprog_prepared.it_ishmed_patient),
coh AS (
  SELECT c71.c_patnr,
         (year(c71.first_dx) - year(CAST(bd.c_birthdate AS TIMESTAMP))) AS age_dx, first_dx
  FROM c71 JOIN bd ON c71.c_patnr=bd.c_patnr)
SELECT count(*) AS n FROM coh WHERE age_dx >= 18;
```

#### A3 — + first diagnosis 2018-2023

*Outcome (Table 2): primary thread — exact; query logged: yes. Ratings DD / GG / consensus: 1 / 1 / 1.*

**Prompt(s) as posed:**

Of these patients (glioblastoma ICD-10 C71.0–C71.9, aged at least 18 at first C71 diagnosis), how many had their first C71 diagnosis between 1 January 2018 and 31 December 2023? Include all diagnoses regardless of verification status.

*[Two aborts; a year-breakdown prompt returned a chart incl. 2024; a single-total prompt returned 1,594 while claiming the 2018–2023 filter. Final prompt:]*

Count the number of distinct adult patients meeting ALL of the following conditions: (1) they have a glioblastoma diagnosis with ICD-10 code C71.0 to C71.9 (any coding slot, regardless of verification status); (2) at the date of their FIRST such C71 diagnosis, they were at least 18 years old (derive age from date of birth and first diagnosis date); (3) the YEAR of that first C71 diagnosis is between 2018 and 2023 inclusive. Apply condition (3) as a filter that excludes any patient whose first C71 diagnosis is in 2024 or later, or before 2018. Return a single total count.

*[Platform asked which columns identify diagnosis, date of birth and diagnosis date.]*

Use the same fields as before: Glioblastoma diagnosis: the ICD-10 code in the Condition resource, i.e. json_extract_scalar(code, '$.coding[0].code'), and also coding[1] and coding[2], matching C71.0 to C71.9. Patient identifier: json_extract_scalar(subject, '$.identifier.value'), joined to the Patient resource via json_extract_scalar(identifier, '$[0].value'). Date of birth: Patient.birthDate, parsed with substr(birthDate, 1, 10). Diagnosis date: Condition.recordedDate; the first diagnosis date is MIN(recordedDate) per patient. Compute age at first diagnosis from birthDate and that MIN(recordedDate). Apply the year filter 2018 to 2023 inclusive on the first diagnosis date, excluding any patient whose first C71 diagnosis is before 2018 or in 2024 or later. Return a single total count.

Platform result: 1587 | Impala reference: 1587 (variant A)

**Generated SQL, primary thread — log entry Jul 31 2026 11:40 · status: no results · rows: 0**

```
SELECT json_extract_scalar(c.subject, '$.identifier.value') AS patient_id, date_trunc('year', cast(c.recordedDate AS timestamp))
AS first_c71_year FROM condition c WHERE (json_extract_scalar(c.code, '$.coding[0].code') LIKE 'C71%' OR
json_extract_scalar(c.code, '$.coding[1].code') LIKE 'C71%' OR json_extract_scalar(c.code, '$.coding[2].code') LIKE 'C71%') AND
cast(date_parse(json_extract_scalar(c.recordedDate, '$'), '%Y-%m-%dT%H:%i:%s') AS date) BETWEEN date '2018-01-01' AND date '2023-
12-31' GROUP BY 1,2
```

**Generated SQL, primary thread — log entry Jul 31 2026 11:43 · status: data available · rows: 7**

```
SELECT
  CAST(year(date_parse(substr(fc.first_c71_date, 1, 10), '%Y-%m-%d')) AS varchar) AS diagnosis_year,
  COUNT(DISTINCT fc.patient_id) AS patient_count
FROM (
  SELECT
    patient_id,
    MIN(recordedDate) AS first_c71_date
  FROM (
    SELECT
      json_extract_scalar(c.subject, '$.identifier.value') AS patient_id,
      c.recordedDate AS recordedDate
    FROM condition c
    WHERE
      json_extract_scalar(c.code, '$.coding[0].code') IN
('C71.0','C71.1','C71.2','C71.3','C71.4','C71.5','C71.6','C71.7','C71.8','C71.9')
      OR json_extract_scalar(c.code, '$.coding[1].code') IN
('C71.0','C71.1','C71.2','C71.3','C71.4','C71.5','C71.6','C71.7','C71.8','C71.9')
      OR json_extract_scalar(c.code, '$.coding[2].code') IN
('C71.0','C71.1','C71.2','C71.3','C71.4','C71.5','C71.6','C71.7','C71.8','C71.9')
  ) q
  GROUP BY patient_id
) fc
JOIN patient p
  ON json_extract_scalar(p.identifier, '$[0].value') = fc.patient_id
WHERE date_diff(
        'year',
        date_parse(p.birthDate, '%Y-%m-%d'),
```

```
        date_parse(substr(fc.first_c71_date, 1, 10), '%Y-%m-%d')
      ) >= 18
GROUP BY CAST(year(date_parse(substr(fc.first_c71_date, 1, 10), '%Y-%m-%d')) AS varchar)
ORDER BY diagnosis_year
```

**Generated SQL, primary thread — log entry Jul 31 2026 11:59 · status: data available · rows: 1**

```
SELECT COUNT(DISTINCT q.patient_id) AS total_patients
FROM (
    SELECT
        dx.patient_id,
        dx.first_recorded_date,
        p.birthDate
    FROM (
        SELECT
            patient_id,
            MIN(recordedDate) AS first_recorded_date
        FROM (
            SELECT
                json_extract_scalar(c.subject, '$.identifier.value') AS patient_id,
                c.recordedDate
            FROM condition c
            WHERE
                json_extract_scalar(c.code, '$.coding[0].code') IN
('C71.0','C71.1','C71.2','C71.3','C71.4','C71.5','C71.6','C71.7','C71.8','C71.9')
                OR json_extract_scalar(c.code, '$.coding[1].code') IN
('C71.0','C71.1','C71.2','C71.3','C71.4','C71.5','C71.6','C71.7','C71.8','C71.9')
                OR json_extract_scalar(c.code, '$.coding[2].code') IN
('C71.0','C71.1','C71.2','C71.3','C71.4','C71.5','C71.6','C71.7','C71.8','C71.9')
        ) matched_conditions
        GROUP BY patient_id
    ) dx
    JOIN patient p
      ON dx.patient_id = json_extract_scalar(p.identifier, '$[0].value')
    WHERE
        year(date_parse(substr(dx.first_recorded_date, 1, 10), '%Y-%m-%d')) BETWEEN 2018 AND 2023
        AND date_diff(
            'year',
            date_parse(substr(p.birthDate, 1, 10), '%Y-%m-%d'),
            date_parse(substr(dx.first_recorded_date, 1, 10), '%Y-%m-%d')
        ) >= 18
) q
```

**Reference SQL (Impala, HDP):**

```
WITH c71 AS (
  SELECT c_patnr, min(c_diagnose_timestamp) AS first_dx
  FROM db_tumpseudoprog_prepared.it_ishmed_diagnose
  WHERE c_diagnose_1 LIKE 'C71%' OR c_diagnose_2 LIKE 'C71%'
  GROUP BY c_patnr),
bd AS (SELECT DISTINCT c_patnr, c_birthdate FROM db_tumpseudoprog_prepared.it_ishmed_patient),
coh AS (
  SELECT c71.c_patnr,
         (year(c71.first_dx) - year(CAST(bd.c_birthdate AS TIMESTAMP))) AS age_dx, first_dx
  FROM c71 JOIN bd ON c71.c_patnr=bd.c_patnr)
SELECT count(*) AS n FROM coh
WHERE age_dx >= 18 AND year(first_dx) BETWEEN 2018 AND 2023;
```

#### A4 — + surgical resection (OPS)

*Outcome (Table 2): primary thread — incorrect but traceable; query logged: yes. Ratings DD / GG / consensus: 2 / 3 / 3.*

**Prompt(s) as posed:**

Take the cohort of adult patients (aged 18 or older at their first C71 diagnosis) with a glioblastoma diagnosis (ICD-10 C71.0 to C71.9, any coding slot, regardless of verification status) whose first C71 diagnosis was in the years 2018 to 2023 inclusive. Of these patients, how many underwent a surgical tumor resection? Identify a surgical resection via the Procedure resource. Return a single total count.

*[Platform offered three resection definitions; the narrow leaf-code set 5-011.0 to 5-016.7 was chosen. A subsequent prefix-match prompt (codes starting with 5-011 … 5-016) led to a clarifying question and an abort.]*

*[v1.2 note: the reference uses prefix matching (regexp ^5-01[1-6]), consistent with B1, V3 and V4, and yields 1275 patients. The platform's 48-code leaf list yields 1248. The 27 missing patients carried codes outside the list: five-digit sub-codes (e.g., 5-016.42, 5-013.40) or codes with the unspecified suffixes .X and .Y (e.g., 5-013.Y, 5-012.X). A patient-level identifier export confirmed the platform's set to be a strict subset of the reference set.]*

Platform result: 1248 | Impala reference: 1275 (prefix matching on 5-011 to 5-016; v1.1 reported 1264 from a regex restricted to one-digit sub-codes, see note below)

#### Generated SQL, primary thread — log entry Jul 31 2026 12:09 • status: data available • rows: 1

```
SELECT COUNT(DISTINCT a.patient_id) AS total_patients
FROM (
    SELECT
        d.patient_id,
        d.first_c71_date
    FROM (
        SELECT
            dx.patient_id,
            MIN(substr(dx.recordedDate, 1, 10)) AS first_c71_date
        FROM (
            SELECT
                json_extract_scalar(c.subject, '$.identifier.value') AS patient_id,
                c.recordedDate AS recordedDate
            FROM condition c
            WHERE
                json_extract_scalar(c.code, '$.coding[0].code') IN
('C71.0','C71.1','C71.2','C71.3','C71.4','C71.5','C71.6','C71.7','C71.8','C71.9')
                OR json_extract_scalar(c.code, '$.coding[1].code') IN
('C71.0','C71.1','C71.2','C71.3','C71.4','C71.5','C71.6','C71.7','C71.8','C71.9')
                OR json_extract_scalar(c.code, '$.coding[2].code') IN
('C71.0','C71.1','C71.2','C71.3','C71.4','C71.5','C71.6','C71.7','C71.8','C71.9')
        ) dx
        GROUP BY dx.patient_id
        HAVING MIN(substr(dx.recordedDate, 1, 10)) BETWEEN '2018-01-01' AND '2023-12-31'
    ) d
    JOIN (
        SELECT
            json_extract_scalar(p.identifier, '$[0].value') AS patient_id,
            p.birthDate AS birthDate
        FROM patient p
    ) pd
        ON d.patient_id = pd.patient_id
    WHERE
        pd.birthDate IS NOT NULL
        AND year(date_parse(d.first_c71_date, '%Y-%m-%d')) - year(date_parse(substr(pd.birthDate, 1, 10), '%Y-%m-%d')) >= 18
) a
JOIN (
    SELECT DISTINCT
        json_extract_scalar(pr.subject, '$.identifier.value') AS patient_id
    FROM procedure pr
    WHERE
        json_extract_scalar(pr.code, '$.coding[0].code') IN (
            '5-011.0','5-011.1','5-011.2','5-011.3','5-011.4','5-011.5','5-011.6','5-011.7',
            '5-012.0','5-012.1','5-012.2','5-012.3','5-012.4','5-012.5','5-012.6','5-012.7',
            '5-013.0','5-013.1','5-013.2','5-013.3','5-013.4','5-013.5','5-013.6','5-013.7',
            '5-014.0','5-014.1','5-014.2','5-014.3','5-014.4','5-014.5','5-014.6','5-014.7',
            '5-015.0','5-015.1','5-015.2','5-015.3','5-015.4','5-015.5','5-015.6','5-015.7',
            '5-016.0','5-016.1','5-016.2','5-016.3','5-016.4','5-016.5','5-016.6','5-016.7'
        )
        OR json_extract_scalar(pr.code, '$.coding[1].code') IN (
            '5-011.0','5-011.1','5-011.2','5-011.3','5-011.4','5-011.5','5-011.6','5-011.7',
            '5-012.0','5-012.1','5-012.2','5-012.3','5-012.4','5-012.5','5-012.6','5-012.7',
            '5-013.0','5-013.1','5-013.2','5-013.3','5-013.4','5-013.5','5-013.6','5-013.7',
            '5-014.0','5-014.1','5-014.2','5-014.3','5-014.4','5-014.5','5-014.6','5-014.7',
            '5-015.0','5-015.1','5-015.2','5-015.3','5-015.4','5-015.5','5-015.6','5-015.7',
            '5-016.0','5-016.1','5-016.2','5-016.3','5-016.4','5-016.5','5-016.6','5-016.7'
        )
        OR json_extract_scalar(pr.code, '$.coding[2].code') IN (
            '5-011.0','5-011.1','5-011.2','5-011.3','5-011.4','5-011.5','5-011.6','5-011.7',
            '5-012.0','5-012.1','5-012.2','5-012.3','5-012.4','5-012.5','5-012.6','5-012.7',
            '5-013.0','5-013.1','5-013.2','5-013.3','5-013.4','5-013.5','5-013.6','5-013.7',
            '5-014.0','5-014.1','5-014.2','5-014.3','5-014.4','5-014.5','5-014.6','5-014.7',
            '5-015.0','5-015.1','5-015.2','5-015.3','5-015.4','5-015.5','5-015.6','5-015.7',
            '5-016.0','5-016.1','5-016.2','5-016.3','5-016.4','5-016.5','5-016.6','5-016.7'
        )
) s
    ON a.patient_id = s.patient_id
```

#### Generated SQL, primary thread — log entry Jul 31 2026 12:17 • status: data available • rows: 1248

```
SELECT DISTINCT a.patient_id
FROM (
    SELECT f.patient_id
    FROM (
        SELECT patient_id, MIN(recordedDate) AS first_c71_recorded_date
        FROM (
            SELECT
                json_extract_scalar(c.subject, '$.identifier.value') AS patient_id,
                c.recordedDate AS recordedDate
            FROM condition c
```

```
            WHERE
                   json_extract_scalar(c.code, '$.coding[0].code') IN ('C71.0', 'C71.1', 'C71.2', 'C71.3', 'C71.4', 'C71.5',
'C71.6', 'C71.7', 'C71.8', 'C71.9')
                OR json_extract_scalar(c.code, '$.coding[1].code') IN ('C71.0', 'C71.1', 'C71.2', 'C71.3', 'C71.4', 'C71.5',
'C71.6', 'C71.7', 'C71.8', 'C71.9')
                OR json_extract_scalar(c.code, '$.coding[2].code') IN ('C71.0', 'C71.1', 'C71.2', 'C71.3', 'C71.4', 'C71.5',
'C71.6', 'C71.7', 'C71.8', 'C71.9')
        ) c71_conditions
        GROUP BY patient_id
    ) f
    JOIN patient p
      ON json_extract_scalar(p.identifier, '$[0].value') = f.patient_id
    WHERE substr(f.first_c71_recorded_date, 1, 10) BETWEEN '2018-01-01' AND '2023-12-31'
      AND (
            year(date_parse(substr(f.first_c71_recorded_date, 1, 10), '%Y-%m-%d'))
            - year(date_parse(substr(p.birthDate, 1, 10), '%Y-%m-%d'))
          ) >= 18
) a
JOIN (
    SELECT DISTINCT
        json_extract_scalar(pr.subject, '$.identifier.value') AS patient_id
    FROM procedure pr
    WHERE
           json_extract_scalar(pr.code, '$.coding[0].code') IN ('5-011.0', '5-011.1', '5-011.2', '5-011.3', '5-011.4', '5-011.5',
'5-011.6', '5-011.7', '5-012.0', '5-012.1', '5-012.2', '5-012.3', '5-012.4', '5-012.5', '5-012.6', '5-012.7', '5-013.0', '5-
013.1', '5-013.2', '5-013.3', '5-013.4', '5-013.5', '5-013.6', '5-013.7', '5-014.0', '5-014.1', '5-014.2', '5-014.3', '5-014.4',
'5-014.5', '5-014.6', '5-014.7', '5-015.0', '5-015.1', '5-015.2', '5-015.3', '5-015.4', '5-015.5', '5-015.6', '5-015.7', '5-
016.0', '5-016.1', '5-016.2', '5-016.3', '5-016.4', '5-016.5', '5-016.6', '5-016.7')
        OR json_extract_scalar(pr.code, '$.coding[1].code') IN ('5-011.0', '5-011.1', '5-011.2', '5-011.3', '5-011.4', '5-011.5',
'5-011.6', '5-011.7', '5-012.0', '5-012.1', '5-012.2', '5-012.3', '5-012.4', '5-012.5', '5-012.6', '5-012.7', '5-013.0', '5-
013.1', '5-013.2', '5-013.3', '5-013.4', '5-013.5', '5-013.6', '5-013.7', '5-014.0', '5-014.1', '5-014.2', '5-014.3', '5-014.4',
'5-014.5', '5-014.6', '5-014.7', '5-015.0', '5-015.1', '5-015.2', '5-015.3', '5-015.4', '5-015.5', '5-015.6', '5-015.7', '5-
016.0', '5-016.1', '5-016.2', '5-016.3', '5-016.4', '5-016.5', '5-016.6', '5-016.7')
        OR json_extract_scalar(pr.code, '$.coding[2].code') IN ('5-011.0', '5-011.1', '5-011.2', '5-011.3', '5-011.4', '5-011.5',
'5-011.6', '5-011.7', '5-012.0', '5-012.1', '5-012.2', '5-012.3', '5-012.4', '5-012.5', '5-012.6', '5-012.7', '5-013.0', '5-
013.1', '5-013.2', '5-013.3', '5-013.4', '5-013.5', '5-013.6', '5-013.7', '5-014.0', '5-014.1', '5-014.2', '5-014.3', '5-014.4',
'5-014.5', '5-014.6', '5-014.7', '5-015.0', '5-015.1', '5-015.2', '5-015.3', '5-015.4', '5-015.5', '5-015.6', '5-015.7', '5-
016.0', '5-016.1', '5-016.2', '5-016.3', '5-016.4', '5-016.5', '5-016.6', '5-016.7')
) r
  ON a.patient_id = r.patient_id
ORDER BY a.patient_id
```

##### Generated SQL, primary thread — log entry Jul 31 2026 12:19 · status: data available · rows: 1248

```
SELECT DISTINCT a.patient_id
FROM (
  SELECT d.patient_id
  FROM (
    SELECT dx.patient_id, MIN(substr(dx.recordedDate,1,10)) AS first_c71_date
    FROM (
      SELECT json_extract_scalar(c.subject,'$.identifier.value') AS patient_id,
             c.recordedDate
      FROM condition c
      WHERE json_extract_scalar(c.code,'$.coding[0].code') IN
('C71.0','C71.1','C71.2','C71.3','C71.4','C71.5','C71.6','C71.7','C71.8','C71.9')
         OR json_extract_scalar(c.code,'$.coding[1].code') IN
('C71.0','C71.1','C71.2','C71.3','C71.4','C71.5','C71.6','C71.7','C71.8','C71.9')
         OR json_extract_scalar(c.code,'$.coding[2].code') IN
('C71.0','C71.1','C71.2','C71.3','C71.4','C71.5','C71.6','C71.7','C71.8','C71.9')
    ) dx
    GROUP BY dx.patient_id
    HAVING MIN(substr(dx.recordedDate,1,10)) BETWEEN '2018-01-01' AND '2023-12-31'
  ) d
  JOIN (
    SELECT json_extract_scalar(p.identifier,'$[0].value') AS patient_id, p.birthDate
    FROM patient p
  ) pd ON d.patient_id = pd.patient_id
  WHERE pd.birthDate IS NOT NULL
    AND year(date_parse(d.first_c71_date,'%Y-%m-%d')) - year(date_parse(substr(pd.birthDate,1,10),'%Y-%m-%d')) >= 18
) a
JOIN (
  SELECT DISTINCT json_extract_scalar(pr.subject,'$.identifier.value') AS patient_id
  FROM procedure pr
  WHERE json_extract_scalar(pr.code,'$.coding[0].code') IN ('5-011.0','5-011.1','5-011.2','5-011.3','5-011.4','5-011.5','5-
011.6','5-011.7','5-012.0','5-012.1','5-012.2','5-012.3','5-012.4','5-012.5','5-012.6','5-012.7','5-013.0','5-013.1','5-013.2','5-
013.3','5-013.4','5-013.5','5-013.6','5-013.7','5-014.0','5-014.1','5-014.2','5-014.3','5-014.4','5-014.5','5-014.6','5-014.7','5-
015.0','5-015.1','5-015.2','5-015.3','5-015.4','5-015.5','5-015.6','5-015.7','5-016.0','5-016.1','5-016.2','5-016.3','5-016.4','5-
016.5','5-016.6','5-016.7')
     OR json_extract_scalar(pr.code,'$.coding[1].code') IN ('5-011.0','5-011.1','5-011.2','5-011.3','5-011.4','5-011.5','5-
011.6','5-011.7','5-012.0','5-012.1','5-012.2','5-012.3','5-012.4','5-012.5','5-012.6','5-012.7','5-013.0','5-013.1','5-013.2','5-
013.3','5-013.4','5-013.5','5-013.6','5-013.7','5-014.0','5-014.1','5-014.2','5-014.3','5-014.4','5-014.5','5-014.6','5-014.7','5-
015.0','5-015.1','5-015.2','5-015.3','5-015.4','5-015.5','5-015.6','5-015.7','5-016.0','5-016.1','5-016.2','5-016.3','5-016.4','5-
```

```
016.5','5-016.6','5-016.7')
     OR json_extract_scalar(pr.code,'$.coding[2].code') IN ('5-011.0','5-011.1','5-011.2','5-011.3','5-011.4','5-011.5','5-
011.6','5-011.7','5-012.0','5-012.1','5-012.2','5-012.3','5-012.4','5-012.5','5-012.6','5-012.7','5-013.0','5-013.1','5-013.2','5-
013.3','5-013.4','5-013.5','5-013.6','5-013.7','5-014.0','5-014.1','5-014.2','5-014.3','5-014.4','5-014.5','5-014.6','5-014.7','5-
015.0','5-015.1','5-015.2','5-015.3','5-015.4','5-015.5','5-015.6','5-015.7','5-016.0','5-016.1','5-016.2','5-016.3','5-016.4','5-
016.5','5-016.6','5-016.7')
) s ON a.patient_id = s.patient_id
```

**Reference SQL (Impala, HDP):**

```
WITH c71 AS (
  SELECT c_patnr, min(c_diagnose_timestamp) AS first_dx
  FROM db_tumpseudoprog_prepared.it_ishmed_diagnose
  WHERE c_diagnose_1 LIKE 'C71%' OR c_diagnose_2 LIKE 'C71%'
  GROUP BY c_patnr),
bd AS (SELECT DISTINCT c_patnr, c_birthdate FROM db_tumpseudoprog_prepared.it_ishmed_patient),
coh AS (
  SELECT c71.c_patnr,
         (year(c71.first_dx) - year(CAST(bd.c_birthdate AS TIMESTAMP))) AS age_dx, first_dx
  FROM c71 JOIN bd ON c71.c_patnr=bd.c_patnr),
res AS (SELECT DISTINCT c_patnr FROM db_tumpseudoprog_prepared.it_ishmed_procedure
        WHERE regexp_like(c_prozedur_code, '^5-01[1-6]'))
SELECT count(*) AS n FROM coh
WHERE age_dx >= 18 AND year(first_dx) BETWEEN 2018 AND 2023
  AND c_patnr IN (SELECT c_patnr FROM res);
```

#### B1 — Sex distribution

*Outcome (Table 2): primary thread — exact; query logged: yes. Ratings DD / GG / consensus: 1 / 1 / 1.*

**Prompt(s) as posed:**

For the final cohort — adult patients (18 or older at their first C71 diagnosis) with a glioblastoma diagnosis (ICD-10 C71.0 to C71.9, any coding slot, regardless of verification status), first diagnosis in 2018 to 2023, who underwent a surgical tumor resection (procedure code starting with 5-011, 5-012, 5-013, 5-014, 5-015, or 5-016) — what is the sex distribution? Give the count of distinct patients per sex.

Platform result: M 741 / F 534 (58.1/41.9%) | Impala reference: M 741 / F 534 (58.1/41.9%)

**Generated SQL, primary thread — log entry Jul 31 2026 12:32 • status: data available • rows: 2**

```
SELECT
    p.gender AS sex,
    COUNT(DISTINCT fc.patient_id) AS patient_count
FROM (
    SELECT DISTINCT a.patient_id
    FROM (
        SELECT
            f.patient_id,
            f.first_dx_date,
            p.gender,
            year(date_parse(substr(f.first_dx_date, 1, 10), '%Y-%m-%d')) - year(date_parse(substr(p.birthDate, 1, 10), '%Y-%m-
%d')) AS age_at_first_dx
        FROM (
            SELECT
                q.patient_id,
                MIN(q.recordedDate) AS first_dx_date
            FROM (
                SELECT
                    json_extract_scalar(c.subject, '$.identifier.value') AS patient_id,
                    c.recordedDate AS recordedDate
                FROM condition c
                WHERE
                    json_extract_scalar(c.code, '$.coding[0].code') IN
('C71.0','C71.1','C71.2','C71.3','C71.4','C71.5','C71.6','C71.7','C71.8','C71.9')
                    OR json_extract_scalar(c.code, '$.coding[1].code') IN
('C71.0','C71.1','C71.2','C71.3','C71.4','C71.5','C71.6','C71.7','C71.8','C71.9')
                    OR json_extract_scalar(c.code, '$.coding[2].code') IN
('C71.0','C71.1','C71.2','C71.3','C71.4','C71.5','C71.6','C71.7','C71.8','C71.9')
            ) q
            GROUP BY q.patient_id
        ) f
        JOIN patient p
          ON json_extract_scalar(p.identifier, '$[0].value') = f.patient_id
        WHERE substr(f.first_dx_date, 1, 10) BETWEEN '2018-01-01' AND '2023-12-31'
          AND p.birthDate IS NOT NULL
          AND year(date_parse(substr(f.first_dx_date, 1, 10), '%Y-%m-%d')) - year(date_parse(substr(p.birthDate, 1, 10), '%Y-%m-
%d')) >= 18
    ) a
    JOIN (
        SELECT DISTINCT
            json_extract_scalar(pr.subject, '$.identifier.value') AS patient_id
```

```
        FROM procedure pr
        WHERE
            json_extract_scalar(pr.code, '$.coding[0].code') LIKE '5-011%'
            OR json_extract_scalar(pr.code, '$.coding[1].code') LIKE '5-011%'
            OR json_extract_scalar(pr.code, '$.coding[2].code') LIKE '5-011%'
            OR json_extract_scalar(pr.code, '$.coding[0].code') LIKE '5-012%'
            OR json_extract_scalar(pr.code, '$.coding[1].code') LIKE '5-012%'
            OR json_extract_scalar(pr.code, '$.coding[2].code') LIKE '5-012%'
            OR json_extract_scalar(pr.code, '$.coding[0].code') LIKE '5-013%'
            OR json_extract_scalar(pr.code, '$.coding[1].code') LIKE '5-013%'
            OR json_extract_scalar(pr.code, '$.coding[2].code') LIKE '5-013%'
            OR json_extract_scalar(pr.code, '$.coding[0].code') LIKE '5-014%'
            OR json_extract_scalar(pr.code, '$.coding[1].code') LIKE '5-014%'
            OR json_extract_scalar(pr.code, '$.coding[2].code') LIKE '5-014%'
            OR json_extract_scalar(pr.code, '$.coding[0].code') LIKE '5-015%'
            OR json_extract_scalar(pr.code, '$.coding[1].code') LIKE '5-015%'
            OR json_extract_scalar(pr.code, '$.coding[2].code') LIKE '5-015%'
            OR json_extract_scalar(pr.code, '$.coding[0].code') LIKE '5-016%'
            OR json_extract_scalar(pr.code, '$.coding[1].code') LIKE '5-016%'
            OR json_extract_scalar(pr.code, '$.coding[2].code') LIKE '5-016%'
    ) pc
      ON a.patient_id = pc.patient_id
) fc
JOIN patient p
  ON json_extract_scalar(p.identifier, '$[0].value') = fc.patient_id
GROUP BY p.gender
ORDER BY p.gender
```

**Reference SQL (Impala, HDP):**

```
WITH c71 AS (
  SELECT c_patnr, min(c_diagnose_timestamp) AS first_dx
  FROM db_tumpseudoprog_prepared.it_ishmed_diagnose
  WHERE c_diagnose_1 LIKE 'C71%' OR c_diagnose_2 LIKE 'C71%'
  GROUP BY c_patnr),
pat AS (
  SELECT DISTINCT c_patnr, c_birthdate, c_gender
  FROM db_tumpseudoprog_prepared.it_ishmed_patient),
coh AS (
  SELECT c71.c_patnr, pat.c_gender,
         (year(c71.first_dx) - year(CAST(pat.c_birthdate AS TIMESTAMP))) AS age_dx,
         year(c71.first_dx) AS dx_year
  FROM c71 JOIN pat ON c71.c_patnr = pat.c_patnr),
res AS (
  SELECT DISTINCT c_patnr
  FROM db_tumpseudoprog_prepared.it_ishmed_procedure
  WHERE regexp_like(c_prozedur_code, '^5-01[1-6]'))
SELECT c_gender, count(DISTINCT coh.c_patnr) AS n
FROM coh JOIN res ON coh.c_patnr = res.c_patnr
WHERE age_dx >= 18 AND dx_year BETWEEN 2018 AND 2023
GROUP BY c_gender
ORDER BY n DESC;
-- v1.2: replaces the v1.1 query, which grouped the whole patient table by sex. Output 16 Oct 2026: M 741, F 534.
```

#### B2 — Mean age at first diagnosis

*Outcome (Table 2): primary thread — exact; query logged: no → yes. Ratings DD / GG / consensus: 1 / 1 / 1. Re-test (new thread): incorrect but traceable; query logged: yes (default filter). Ratings DD / GG / consensus: 3 / 2 / 3.*

**Prompt(s) as posed:**

For adult patients with a glioblastoma diagnosis (ICD-10 C71.0 to C71.9, any coding slot, regardless of verification status), what is the mean age at their first C71 diagnosis? Compute age from date of birth and first diagnosis date. Give one mean value.

*[Answer 64.9 years, no query logged. After three aborts, the final prompt:]*

For the distinct adult patients with a C71 diagnosis (any subcode), summarize age at first diagnosis

Platform result: 55.09 (n=1594) | Impala reference: 55.12 (n=1594)

**Generated SQL, primary thread — log entry Jul 31 2026 12:44 · status: data available · rows: 1936**

```
SELECT
  cohort.patient_id,
  cohort.first_dx_date,
  p.birthDate AS birth_date,
  year(date_parse(cohort.first_dx_date, '%Y-%m-%d')) - year(date_parse(p.birthDate, '%Y-%m-%d')) AS age
FROM (
  SELECT
    dx.patient_id,
    substr(MIN(dx.recorded_ts), 1, 10) AS first_dx_date
  FROM (
```

```
    SELECT
      json_extract_scalar(c.subject, '$.identifier.value') AS patient_id,
      c.recordedDate AS recorded_ts
    FROM condition c
    WHERE
      json_extract_scalar(c.code, '$.coding[0].code') IN
('C71.0','C71.1','C71.2','C71.3','C71.4','C71.5','C71.6','C71.7','C71.8','C71.9')
      OR json_extract_scalar(c.code, '$.coding[1].code') IN
('C71.0','C71.1','C71.2','C71.3','C71.4','C71.5','C71.6','C71.7','C71.8','C71.9')
      OR json_extract_scalar(c.code, '$.coding[2].code') IN
('C71.0','C71.1','C71.2','C71.3','C71.4','C71.5','C71.6','C71.7','C71.8','C71.9')
  ) dx
  WHERE dx.patient_id IS NOT NULL
    AND dx.recorded_ts IS NOT NULL
  GROUP BY dx.patient_id
) cohort
JOIN patient p
  ON cohort.patient_id = json_extract_scalar(p.identifier, '$[0].value')
WHERE p.birthDate IS NOT NULL
  AND cohort.first_dx_date IS NOT NULL
  AND year(date_parse(cohort.first_dx_date, '%Y-%m-%d')) - year(date_parse(p.birthDate, '%Y-%m-%d')) >= 18
```

Re-test (new thread, 1 Sep 2026): platform result 53.2 y, n=113

*[Re-test classification: the logged SQL carries verificationStatus = 'G' although the prompt said "regardless of verification status". Rebuilding the reference with c_gewissheit = 'G' reproduces n = 113 (mean 54.1 y by calendar-year age; the platform used completed years). Classified as incorrect but traceable.]*

**Generated SQL, re-test — log entry Sep 01 2026 14:11 · status: data available · rows: 1**

```
SELECT
  COUNT(DISTINCT patient_id) AS n_patients,
  AVG(age_at_first_dx) AS mean_age_at_first_dx
FROM (
  SELECT
    patient_id,
    date_diff('year', birth_dt, first_dx_dt) AS age_at_first_dx
  FROM (
    SELECT
      dx.patient_id,
      p.birth_dt,
      dx.first_dx_dt
    FROM (
      SELECT
        patient_id,
        MIN(recorded_dt) AS first_dx_dt
      FROM (
        SELECT
          json_extract_scalar(c.subject, '$.identifier.value') AS patient_id,
          date_parse(substr(c.recordedDate, 1, 10), '%Y-%m-%d') AS recorded_dt
        FROM condition c
        WHERE json_extract_scalar(c.verificationStatus, '$.text') = 'G'
          AND (
            json_extract_scalar(c.code, '$.coding[0].code') LIKE 'C71%'
            OR json_extract_scalar(c.code, '$.coding[1].code') LIKE 'C71%'
            OR json_extract_scalar(c.code, '$.coding[2].code') LIKE 'C71%'
          )
      ) cond
      WHERE recorded_dt IS NOT NULL
      GROUP BY patient_id
    ) dx
    JOIN (
      SELECT
        json_extract_scalar(p.identifier, '$[0].value') AS patient_id,
        date_parse(substr(p.birthDate, 1, 10), '%Y-%m-%d') AS birth_dt
      FROM patient p
      WHERE p.birthDate IS NOT NULL
    ) p
      ON dx.patient_id = p.patient_id
  ) joined_data
  WHERE birth_dt IS NOT NULL
    AND date_diff('year', birth_dt, first_dx_dt) >= 18
) adults
```

**Reference SQL (Impala, HDP):**

```
WITH c71 AS (
  SELECT c_patnr, min(c_diagnose_timestamp) AS first_dx
  FROM db_tumpseudoprog_prepared.it_ishmed_diagnose
  WHERE c_diagnose_1 LIKE 'C71%' OR c_diagnose_2 LIKE 'C71%'
  GROUP BY c_patnr),
bd AS (SELECT DISTINCT c_patnr, c_birthdate FROM db_tumpseudoprog_prepared.it_ishmed_patient),
```

```
coh AS (
  SELECT c71.c_patnr,
         (year(c71.first_dx) - year(CAST(bd.c_birthdate AS TIMESTAMP))) AS age_dx, first_dx
  FROM c71 JOIN bd ON c71.c_patnr=bd.c_patnr)
SELECT round(avg(age_dx),1) AS mean_age FROM coh;
```

#### B3 — Mortality (deceased patients)

##### *Outcome (Table 2): primary thread — exact; query logged: yes. Ratings DD / GG / consensus: 1 / 1 / 1.*

**Prompt(s) as posed:**

Among adult patients (18 or older at first C71 diagnosis) with a glioblastoma diagnosis (ICD-10 C71.0 to C71.9 in any coding slot, regardless of verification status), how many are deceased? Use all such patients with no further filtering. State how many patients are included in total and how many of them are deceased.

Platform result: 101 / 1594 (6.3%) | Impala reference: 101 / 1594 (6.3%)

**Generated SQL, primary thread — log entry Jul 31 2026 13:33 · status: data available · rows: 1**

```
SELECT
  COUNT(DISTINCT patient_id) AS total_included_adult_glioblastoma_patients,
  COUNT(DISTINCT CASE WHEN deceased_flag = 1 THEN patient_id END) AS number_deceased
FROM (
  SELECT
    dx.patient_id,
    CASE WHEN p.deceasedDateTime IS NOT NULL THEN 1 ELSE 0 END AS deceased_flag,
    date_diff(
      'year',
      date_parse(substr(p.birthDate, 1, 10), '%Y-%m-%d'),
      date_parse(substr(dx.first_c71_recorded_date, 1, 10), '%Y-%m-%d')
    ) AS age_at_first_c71_diagnosis
  FROM (
    SELECT
      json_extract_scalar(c.subject, '$.identifier.value') AS patient_id,
      MIN(c.recordedDate) AS first_c71_recorded_date
    FROM condition c
    WHERE
         json_extract_scalar(c.code, '$.coding[0].code') IN
('C71.0','C71.1','C71.2','C71.3','C71.4','C71.5','C71.6','C71.7','C71.8','C71.9')
      OR json_extract_scalar(c.code, '$.coding[1].code') IN
('C71.0','C71.1','C71.2','C71.3','C71.4','C71.5','C71.6','C71.7','C71.8','C71.9')
      OR json_extract_scalar(c.code, '$.coding[2].code') IN
('C71.0','C71.1','C71.2','C71.3','C71.4','C71.5','C71.6','C71.7','C71.8','C71.9')
    GROUP BY 1
  ) dx
  JOIN patient p
    ON dx.patient_id = json_extract_scalar(p.identifier, '$[0].value')
  WHERE p.birthDate IS NOT NULL
) adult_cohort
WHERE age_at_first_c71_diagnosis >= 18
```

**Reference SQL (Impala, HDP):**

```
SELECT count(DISTINCT c_patnr) AS n FROM db_tumpseudoprog_prepared.it_ishmed_patient
WHERE c_datetimeofdeath IS NOT NULL;
```

#### B4 — Most frequent OPS codes

##### *Outcome (Table 2): primary thread — exact; query logged: yes. Ratings DD / GG / consensus: 1 / 1 / 1.*

**Prompt(s) as posed:**

Among adult patients (18 or older at first C71 diagnosis) with a glioblastoma diagnosis (ICD-10 C71.0 to C71.9 in any coding slot, regardless of verification status), what are the 15 most frequent surgical procedure (OPS) codes recorded for these patients? List each procedure code with the number of distinct patients who have it. Order by patient count descending.

Platform result: Top-15 identical | Impala reference: Top-15 identical

**Generated SQL, primary thread — log entry Jul 31 2026 13:36 · status: data available · rows: 15**

```
SELECT
    ops_code,
    patient_count
FROM (
    SELECT
        ops_code,
        COUNT(DISTINCT patient_id) AS patient_count
    FROM (
        SELECT
```

```
            ac.patient_id,
            COALESCE(
                json_extract_scalar(pr.code, '$.coding[0].code'),
                json_extract_scalar(pr.code, '$.coding[1].code'),
                json_extract_scalar(pr.code, '$.coding[2].code')
            ) AS ops_code
        FROM (
            SELECT
                f.patient_id,
                f.first_dx_date,
                year(date_parse(f.first_dx_date, '%Y-%m-%d')) - year(date_parse(substr(p.birthDate, 1, 10), '%Y-%m-%d')) AS
age_at_first_dx
            FROM (
                SELECT
                    patient_id,
                    MIN(dx_date) AS first_dx_date
                FROM (
                    SELECT
                        json_extract_scalar(c.subject, '$.identifier.value') AS patient_id,
                        substr(c.recordedDate, 1, 10) AS dx_date
                    FROM condition c
                    WHERE
                        json_extract_scalar(c.code, '$.coding[0].code') IN
('C71.0','C71.1','C71.2','C71.3','C71.4','C71.5','C71.6','C71.7','C71.8','C71.9')
                        OR json_extract_scalar(c.code, '$.coding[1].code') IN
('C71.0','C71.1','C71.2','C71.3','C71.4','C71.5','C71.6','C71.7','C71.8','C71.9')
                        OR json_extract_scalar(c.code, '$.coding[2].code') IN
('C71.0','C71.1','C71.2','C71.3','C71.4','C71.5','C71.6','C71.7','C71.8','C71.9')
                ) qualifying_dx
                WHERE dx_date IS NOT NULL
                GROUP BY patient_id
            ) f
            JOIN patient p
                ON json_extract_scalar(p.identifier, '$[0].value') = f.patient_id
            WHERE p.birthDate IS NOT NULL
              AND f.first_dx_date IS NOT NULL
              AND year(date_parse(f.first_dx_date, '%Y-%m-%d')) - year(date_parse(substr(p.birthDate, 1, 10), '%Y-%m-%d')) >= 18
        ) ac
        JOIN procedure pr
            ON json_extract_scalar(pr.subject, '$.identifier.value') = ac.patient_id
        WHERE COALESCE(
                json_extract_scalar(pr.code, '$.coding[0].code'),
                json_extract_scalar(pr.code, '$.coding[1].code'),
                json_extract_scalar(pr.code, '$.coding[2].code')
              ) IS NOT NULL
    ) ops_rows
    GROUP BY ops_code
) ranked_ops
ORDER BY patient_count DESC, ops_code
LIMIT 15
```

**Reference SQL (Impala, HDP):**

```
WITH c71 AS (
  SELECT c_patnr, min(c_diagnose_timestamp) AS first_dx
  FROM db_tumpseudoprog_prepared.it_ishmed_diagnose
  WHERE c_diagnose_1 LIKE 'C71%' OR c_diagnose_2 LIKE 'C71%'
  GROUP BY c_patnr),
bd AS (SELECT DISTINCT c_patnr, c_birthdate FROM db_tumpseudoprog_prepared.it_ishmed_patient),
coh AS (
  SELECT c71.c_patnr
  FROM c71 JOIN bd ON c71.c_patnr = bd.c_patnr
  WHERE (year(c71.first_dx) - year(CAST(bd.c_birthdate AS TIMESTAMP))) >= 18)
SELECT p.c_prozedur_code, count(DISTINCT p.c_patnr) AS n_patients
FROM db_tumpseudoprog_prepared.it_ishmed_procedure p
JOIN coh ON p.c_patnr = coh.c_patnr
GROUP BY p.c_prozedur_code
ORDER BY n_patients DESC
LIMIT 15;
-- v1.2: replaces the v1.1 query, which counted procedure rows over all patients. Output 11 Oct 2026: 15/15 codes and counts
identical to the platform (3-820 1415, 5-984 1275, 8-930 1233, ...).
```

#### V1 — Tumor location distribution (C71.0-C71.9 subcodes)

*Outcome (Table 2): primary thread — exact; query logged: yes. Ratings DD / GG / consensus: 1 / 1 / 1.*

**Prompt(s) as posed:**

Among adult patients (18 or older at first C71 diagnosis) with a glioblastoma diagnosis (ICD-10 C71.0 to C71.9 in any coding slot, regardless of verification status), how are patients distributed across the C71 subcodes (C71.0 to C71.9)? Count distinct patients per subcode; a patient may appear under more than one subcode. Order by patient count descending.

Platform result: 10/10 subcodes identical (C71.1 457 … C71.5 51); sum 2208 | Impala reference: 10/10 identical; sum 2208 (>1594, multi-location)

**Generated SQL, primary thread — log entry Aug 25 2026 22:26 · status: data available · rows: 10**

```
SELECT
  subcode,
  COUNT(DISTINCT patient_id) AS patient_count
FROM (
  SELECT DISTINCT
    patient_id,
    matched_code AS subcode
  FROM (
    SELECT
      patient_id,
      matched_code,
      first_c71_date,
      birth_date,
      date_diff(
        'year',
        date_parse(substr(birth_date, 1, 10), '%Y-%m-%d'),
        date_parse(substr(first_c71_date, 1, 10), '%Y-%m-%d')
      ) AS age_at_first_c71
    FROM (
      SELECT
        d.patient_id,
        d.matched_code,
        f.first_c71_date,
        p.birthDate AS birth_date
      FROM (
        SELECT
          json_extract_scalar(c.subject, '$.identifier.value') AS patient_id,
          CASE
            WHEN json_extract_scalar(c.code, '$.coding[0].code') IN
('C71.0','C71.1','C71.2','C71.3','C71.4','C71.5','C71.6','C71.7','C71.8','C71.9')
              THEN json_extract_scalar(c.code, '$.coding[0].code')
            WHEN json_extract_scalar(c.code, '$.coding[1].code') IN
('C71.0','C71.1','C71.2','C71.3','C71.4','C71.5','C71.6','C71.7','C71.8','C71.9')
              THEN json_extract_scalar(c.code, '$.coding[1].code')
            WHEN json_extract_scalar(c.code, '$.coding[2].code') IN
('C71.0','C71.1','C71.2','C71.3','C71.4','C71.5','C71.6','C71.7','C71.8','C71.9')
              THEN json_extract_scalar(c.code, '$.coding[2].code')
          END AS matched_code
        FROM condition c
        WHERE json_extract_scalar(c.code, '$.coding[0].code') IN
('C71.0','C71.1','C71.2','C71.3','C71.4','C71.5','C71.6','C71.7','C71.8','C71.9')
           OR json_extract_scalar(c.code, '$.coding[1].code') IN
('C71.0','C71.1','C71.2','C71.3','C71.4','C71.5','C71.6','C71.7','C71.8','C71.9')
           OR json_extract_scalar(c.code, '$.coding[2].code') IN
('C71.0','C71.1','C71.2','C71.3','C71.4','C71.5','C71.6','C71.7','C71.8','C71.9')
      ) d
      JOIN (
        SELECT
          patient_id,
          MIN(recorded_dt) AS first_c71_date
        FROM (
          SELECT
            json_extract_scalar(c.subject, '$.identifier.value') AS patient_id,
            c.recordedDate AS recorded_dt
          FROM condition c
          WHERE json_extract_scalar(c.code, '$.coding[0].code') IN
('C71.0','C71.1','C71.2','C71.3','C71.4','C71.5','C71.6','C71.7','C71.8','C71.9')
             OR json_extract_scalar(c.code, '$.coding[1].code') IN
('C71.0','C71.1','C71.2','C71.3','C71.4','C71.5','C71.6','C71.7','C71.8','C71.9')
             OR json_extract_scalar(c.code, '$.coding[2].code') IN
('C71.0','C71.1','C71.2','C71.3','C71.4','C71.5','C71.6','C71.7','C71.8','C71.9')
        ) firsts
        GROUP BY patient_id
      ) f
        ON d.patient_id = f.patient_id
      JOIN patient p
        ON d.patient_id = json_extract_scalar(p.identifier, '$[0].value')
      WHERE d.matched_code IS NOT NULL
        AND p.birthDate IS NOT NULL
        AND f.first_c71_date IS NOT NULL
    ) joined_data
  ) aged
```

```
  WHERE age_at_first_c71 >= 18
) final_counts
GROUP BY subcode
ORDER BY subcode
```

**Reference SQL (Impala, HDP):**

```
WITH c71 AS (
  SELECT c_patnr, min(c_diagnose_timestamp) AS first_dx
  FROM db_tumpseudoprog_prepared.it_ishmed_diagnose
  WHERE c_diagnose_1 LIKE 'C71%' OR c_diagnose_2 LIKE 'C71%'
  GROUP BY c_patnr),
bd AS (SELECT DISTINCT c_patnr, c_birthdate FROM db_tumpseudoprog_prepared.it_ishmed_patient),
coh AS (
  SELECT c71.c_patnr, c71.first_dx,
         (year(c71.first_dx) - year(CAST(bd.c_birthdate AS TIMESTAMP))) AS age_dx
  FROM c71 JOIN bd ON c71.c_patnr = bd.c_patnr
  WHERE (year(c71.first_dx) - year(CAST(bd.c_birthdate AS TIMESTAMP))) >= 18),
sub AS (
  SELECT DISTINCT d.c_patnr, substr(CASE WHEN d.c_diagnose_1 LIKE 'C71%' THEN d.c_diagnose_1 ELSE d.c_diagnose_2 END,1,5) AS
subcode
  FROM db_tumpseudoprog_prepared.it_ishmed_diagnose d
  WHERE d.c_diagnose_1 LIKE 'C71%' OR d.c_diagnose_2 LIKE 'C71%')
SELECT s.subcode, count(DISTINCT s.c_patnr) AS n_patients
FROM sub s JOIN coh ON s.c_patnr = coh.c_patnr
GROUP BY s.subcode ORDER BY n_patients DESC;
```

#### V2 — Biopsy vs. resection split

*Outcome (Table 2): primary thread — incorrect and unlogged; query logged: no. Ratings DD / GG / consensus: 4 / 4 / 4. Re-test (new thread): not answered; query logged: —. Ratings DD / GG / consensus: 5 / 5 / 5.*

**Prompt(s) as posed:**

Among the adult C71 cohort, how many patients underwent (a) a surgical tumor resection (procedure code starting with 5-011 to 5-016) and how many underwent (b) a brain biopsy only (biopsy procedure code, no resection code)? Report both counts of distinct patients.

Platform result: resection 1275; biopsy-only 117 | Impala reference: resection 1279; biopsy-only 258

**Generated SQL, primary thread: no query logged.**

Re-test (new thread, 1 Sep 2026): platform result explained abstention

**Generated SQL, re-test: no query logged.**

**Reference SQL (Impala, HDP):**

```
WITH c71 AS (
  SELECT c_patnr, min(c_diagnose_timestamp) AS first_dx
  FROM db_tumpseudoprog_prepared.it_ishmed_diagnose
  WHERE c_diagnose_1 LIKE 'C71%' OR c_diagnose_2 LIKE 'C71%'
  GROUP BY c_patnr),
bd AS (SELECT DISTINCT c_patnr, c_birthdate FROM db_tumpseudoprog_prepared.it_ishmed_patient),
coh AS (
  SELECT c71.c_patnr, c71.first_dx,
         (year(c71.first_dx) - year(CAST(bd.c_birthdate AS TIMESTAMP))) AS age_dx
  FROM c71 JOIN bd ON c71.c_patnr = bd.c_patnr
  WHERE (year(c71.first_dx) - year(CAST(bd.c_birthdate AS TIMESTAMP))) >= 18),
res AS (SELECT DISTINCT c_patnr FROM db_tumpseudoprog_prepared.it_ishmed_procedure
        WHERE regexp_like(c_prozedur_code,'^5-01[1-6]')),
bio AS (SELECT DISTINCT c_patnr FROM db_tumpseudoprog_prepared.it_ishmed_procedure
        WHERE c_prozedur_code LIKE '1-51%')  -- biopsy code set TO CONFIRM with Jawed
SELECT
  count(DISTINCT CASE WHEN coh.c_patnr IN (SELECT c_patnr FROM res) THEN coh.c_patnr END) AS n_resection,
  count(DISTINCT CASE WHEN coh.c_patnr IN (SELECT c_patnr FROM bio)
                       AND coh.c_patnr NOT IN (SELECT c_patnr FROM res) THEN coh.c_patnr END) AS n_biopsy_only
FROM coh;
```

#### V3 — Median days from first diagnosis to first resection

*Outcome (Table 2): primary thread — exact; query logged: yes. Ratings DD / GG / consensus: 1 / 1 / 1.*

**Prompt(s) as posed:**

Among adult C71 patients who underwent a surgical tumor resection (code starting 5-011 to 5-016), what is the median number of days between the first C71 diagnosis and the first resection procedure? State how many patients are included.

Platform result: median 0 d; 49.5% on / 26.1% before / 24.4% after (n=1279) | Impala reference: median 0 d; 49.5% / 26.1% / 24.4% (n=1279)

##### Generated SQL, primary thread — log entry Aug 25 2026 22:15 • status: data available • rows: 1610

```
SELECT
  d.patient_id,
  d.first_dx_date,
  r.first_resection_date,
  date_diff(
    'day',
    date_parse(substr(d.first_dx_date, 1, 10), '%Y-%m-%d'),
    date_parse(substr(r.first_resection_date, 1, 10), '%Y-%m-%d')
  ) AS days_to_resection,
  date_diff(
    'year',
    date_parse(substr(p.birthDate, 1, 10), '%Y-%m-%d'),
    date_parse(substr(d.first_dx_date, 1, 10), '%Y-%m-%d')
  ) AS age_at_dx
FROM (
  SELECT
    patient_id,
    MIN(recordedDate) AS first_dx_date
  FROM (
    SELECT
      json_extract_scalar(c.subject, '$.identifier.value') AS patient_id,
      c.recordedDate,
      json_extract_scalar(c.code, '$.coding[0].code') AS code0,
      json_extract_scalar(c.code, '$.coding[1].code') AS code1,
      json_extract_scalar(c.code, '$.coding[2].code') AS code2
    FROM condition c
    WHERE c.recordedDate IS NOT NULL
  ) dx
  WHERE code0 IN ('C71.0','C71.1','C71.2','C71.3','C71.4','C71.5','C71.6','C71.7','C71.8','C71.9')
     OR code1 IN ('C71.0','C71.1','C71.2','C71.3','C71.4','C71.5','C71.6','C71.7','C71.8','C71.9')
     OR code2 IN ('C71.0','C71.1','C71.2','C71.3','C71.4','C71.5','C71.6','C71.7','C71.8','C71.9')
  GROUP BY patient_id
) d
JOIN (
  SELECT
    patient_id,
    MIN(proc_start) AS first_resection_date
  FROM (
    SELECT
      json_extract_scalar(pr.subject, '$.identifier.value') AS patient_id,
      json_extract_scalar(pr.performedPeriod, '$.start') AS proc_start,
      json_extract_scalar(pr.code, '$.coding[0].code') AS code0,
      json_extract_scalar(pr.code, '$.coding[1].code') AS code1,
      json_extract_scalar(pr.code, '$.coding[2].code') AS code2
    FROM procedure pr
    WHERE json_extract_scalar(pr.performedPeriod, '$.start') IS NOT NULL
  ) px
  WHERE code0 LIKE '5-011%' OR code1 LIKE '5-011%' OR code2 LIKE '5-011%'
     OR code0 LIKE '5-012%' OR code1 LIKE '5-012%' OR code2 LIKE '5-012%'
     OR code0 LIKE '5-013%' OR code1 LIKE '5-013%' OR code2 LIKE '5-013%'
     OR code0 LIKE '5-014%' OR code1 LIKE '5-014%' OR code2 LIKE '5-014%'
     OR code0 LIKE '5-015%' OR code1 LIKE '5-015%' OR code2 LIKE '5-015%'
     OR code0 LIKE '5-016%' OR code1 LIKE '5-016%' OR code2 LIKE '5-016%'
  GROUP BY patient_id
) r
  ON d.patient_id = r.patient_id
JOIN patient p
  ON d.patient_id = json_extract_scalar(p.identifier, '$[0].value')
WHERE p.birthDate IS NOT NULL
  AND date_diff(
        'year',
        date_parse(substr(p.birthDate, 1, 10), '%Y-%m-%d'),
        date_parse(substr(d.first_dx_date, 1, 10), '%Y-%m-%d')
      ) >= 18
```

##### Reference SQL (Impala, HDP):

```
WITH c71 AS (
  SELECT c_patnr, min(c_diagnose_timestamp) AS first_dx
  FROM db_tumpseudoprog_prepared.it_ishmed_diagnose
  WHERE c_diagnose_1 LIKE 'C71%' OR c_diagnose_2 LIKE 'C71%'
  GROUP BY c_patnr),
bd AS (SELECT DISTINCT c_patnr, c_birthdate FROM db_tumpseudoprog_prepared.it_ishmed_patient),
coh AS (
  SELECT c71.c_patnr, c71.first_dx,
         (year(c71.first_dx) - year(CAST(bd.c_birthdate AS TIMESTAMP))) AS age_dx
  FROM c71 JOIN bd ON c71.c_patnr = bd.c_patnr
  WHERE (year(c71.first_dx) - year(CAST(bd.c_birthdate AS TIMESTAMP))) >= 18),
first_res AS (
  SELECT c_patnr, min(c_prozedur_begin) AS first_res_ts
```

```
  FROM db_tumpseudoprog_prepared.it_ishmed_procedure
  WHERE regexp_like(c_prozedur_code,'^5-01[1-6]')
  GROUP BY c_patnr)
SELECT coh.c_patnr,
       datediff(CAST(fr.first_res_ts AS TIMESTAMP), CAST(coh.first_dx AS TIMESTAMP)) AS days_dx_to_res
FROM coh JOIN first_res fr ON coh.c_patnr = fr.c_patnr;
-- EXPORT per-patient rows, compute median offline (desserd lacks percentile_approx privilege)
```

#### V4 — Re-resection rate (patients with >= 2 resections)

*Outcome (Table 2): primary thread — incorrect but traceable; query logged: yes (default filter). Ratings DD / GG / consensus: 3 / 3 / 3. Re-test (new thread): incorrect but traceable; query logged: yes (default filter + date restriction). Ratings DD / GG / consensus: 3 / 3 / 3.*

**Prompt(s) as posed:**

Among adult C71 patients with at least one surgical tumor resection (code starting 5-011 to 5-016), how many had two or more resections on distinct dates? Report the count and the share of all resected patients.

Platform result: re-resected 34; total resected 95; 35.8% | Impala reference: re-resected 220; total resected 1279; 17.2%

**Generated SQL, primary thread — log entry Aug 25 2026 22:41 • status: data available • rows: 1**

```
SELECT
  COUNT(DISTINCT CASE WHEN proc_date_count >= 2 THEN patient_id END) AS numerator,
  COUNT(DISTINCT patient_id) AS denominator
FROM (
  SELECT
    adult_dx.patient_id,
    COUNT(DISTINCT adult_dx.proc_date) AS proc_date_count
  FROM (
    SELECT
      dx.patient_id,
      dx.first_dx_date,
      CAST(date_diff('year', date_parse(substr(p.birthDate, 1, 10), '%Y-%m-%d'), date_parse(substr(dx.first_dx_date, 1, 10), '%Y-
%m-%d')) AS INTEGER) AS age_at_dx,
      proc.proc_date
    FROM (
      SELECT
        patient_id,
        MIN(recorded_date) AS first_dx_date
      FROM (
        SELECT
          json_extract_scalar(c.subject, '$.identifier.value') AS patient_id,
          c.recordedDate AS recorded_date
        FROM condition c
        WHERE (
              json_extract_scalar(c.code, '$.coding[0].code') IN
('C71.0','C71.1','C71.2','C71.3','C71.4','C71.5','C71.6','C71.7','C71.8','C71.9')
           OR json_extract_scalar(c.code, '$.coding[1].code') IN
('C71.0','C71.1','C71.2','C71.3','C71.4','C71.5','C71.6','C71.7','C71.8','C71.9')
           OR json_extract_scalar(c.code, '$.coding[2].code') IN
('C71.0','C71.1','C71.2','C71.3','C71.4','C71.5','C71.6','C71.7','C71.8','C71.9')
        )
          AND json_extract_scalar(c.verificationStatus, '$.text') = 'G'
          AND c.recordedDate IS NOT NULL
      ) dx_codes
      GROUP BY patient_id
    ) dx
    JOIN patient p
      ON json_extract_scalar(p.identifier, '$[0].value') = dx.patient_id
    JOIN (
      SELECT
        json_extract_scalar(pr.subject, '$.identifier.value') AS patient_id,
        substr(json_extract_scalar(pr.performedPeriod, '$.start'), 1, 10) AS proc_date
      FROM procedure pr
      WHERE (
            json_extract_scalar(pr.code, '$.coding[0].code') LIKE '5-011%'
         OR json_extract_scalar(pr.code, '$.coding[1].code') LIKE '5-011%'
         OR json_extract_scalar(pr.code, '$.coding[2].code') LIKE '5-011%'
         OR json_extract_scalar(pr.code, '$.coding[0].code') LIKE '5-012%'
         OR json_extract_scalar(pr.code, '$.coding[1].code') LIKE '5-012%'
         OR json_extract_scalar(pr.code, '$.coding[2].code') LIKE '5-012%'
         OR json_extract_scalar(pr.code, '$.coding[0].code') LIKE '5-013%'
         OR json_extract_scalar(pr.code, '$.coding[1].code') LIKE '5-013%'
         OR json_extract_scalar(pr.code, '$.coding[2].code') LIKE '5-013%'
         OR json_extract_scalar(pr.code, '$.coding[0].code') LIKE '5-014%'
         OR json_extract_scalar(pr.code, '$.coding[1].code') LIKE '5-014%'
         OR json_extract_scalar(pr.code, '$.coding[2].code') LIKE '5-014%'
         OR json_extract_scalar(pr.code, '$.coding[0].code') LIKE '5-015%'
```

```
          OR json_extract_scalar(pr.code, '$.coding[1].code') LIKE '5-015%'
          OR json_extract_scalar(pr.code, '$.coding[2].code') LIKE '5-015%'
          OR json_extract_scalar(pr.code, '$.coding[0].code') LIKE '5-016%'
          OR json_extract_scalar(pr.code, '$.coding[1].code') LIKE '5-016%'
          OR json_extract_scalar(pr.code, '$.coding[2].code') LIKE '5-016%'
      )
        AND json_extract_scalar(pr.performedPeriod, '$.start') IS NOT NULL
    ) proc
      ON proc.patient_id = dx.patient_id
    WHERE p.birthDate IS NOT NULL
      AND date_diff('year', date_parse(substr(p.birthDate, 1, 10), '%Y-%m-%d'), date_parse(substr(dx.first_dx_date, 1, 10), '%Y-
%m-%d')) >= 18
  ) adult_dx
  GROUP BY adult_dx.patient_id
) counted
```

Re-test (new thread, 1 Sep 2026): platform result 13 / 83 (15.66%)

##### Generated SQL, re-test — log entry Sep 01 2026 14:38 · status: data available · rows: 1

```
SELECT
  COUNT(DISTINCT CASE WHEN proc_days >= 2 THEN patient_id END) AS numerator,
  COUNT(DISTINCT patient_id) AS denominator
FROM (
  SELECT
    dx.patient_id,
    COUNT(DISTINCT CAST(substr(proc.proc_start, 1, 10) AS varchar)) AS proc_days
  FROM (
    SELECT
      patient_id,
      MIN(recorded_date) AS first_dx_date
    FROM (
      SELECT
        json_extract_scalar(c.subject, '$.identifier.value') AS patient_id,
        c.recordedDate AS recorded_date,
        json_extract_scalar(c.code, '$.coding[0].code') AS code0,
        json_extract_scalar(c.code, '$.coding[1].code') AS code1,
        json_extract_scalar(c.code, '$.coding[2].code') AS code2,
        json_extract_scalar(c.verificationStatus, '$.text') AS verification_status
      FROM condition c
    ) dx0
    WHERE verification_status = 'G'
      AND (
        code0 IN ('C71.0','C71.1','C71.2','C71.3','C71.4','C71.5','C71.6','C71.7','C71.8','C71.9')
        OR code1 IN ('C71.0','C71.1','C71.2','C71.3','C71.4','C71.5','C71.6','C71.7','C71.8','C71.9')
        OR code2 IN ('C71.0','C71.1','C71.2','C71.3','C71.4','C71.5','C71.6','C71.7','C71.8','C71.9')
      )
    GROUP BY patient_id
  ) dx
  JOIN (
    SELECT
      json_extract_scalar(p.identifier, '$[0].value') AS patient_id,
      p.birthDate AS birth_date
    FROM patient p
  ) pat
    ON pat.patient_id = dx.patient_id
  JOIN (
    SELECT
      json_extract_scalar(pr.subject, '$.identifier.value') AS patient_id,
      json_extract_scalar(pr.performedPeriod, '$.start') AS proc_start,
      json_extract_scalar(pr.code, '$.coding[0].code') AS code0,
      json_extract_scalar(pr.code, '$.coding[1].code') AS code1,
      json_extract_scalar(pr.code, '$.coding[2].code') AS code2
    FROM procedure pr
  ) proc
    ON proc.patient_id = dx.patient_id
  WHERE date_diff(
          'year',
          date_parse(substr(pat.birth_date, 1, 10), '%Y-%m-%d'),
          date_parse(substr(dx.first_dx_date, 1, 10), '%Y-%m-%d')
        ) >= 18
    AND proc.proc_start IS NOT NULL
    AND date_parse(substr(proc.proc_start, 1, 10), '%Y-%m-%d') >= date_parse(substr(dx.first_dx_date, 1, 10), '%Y-%m-%d')
    AND (
      code0 LIKE '5-011%' OR code0 LIKE '5-012%' OR code0 LIKE '5-013%' OR code0 LIKE '5-014%' OR code0 LIKE '5-015%' OR code0
LIKE '5-016%'
      OR code1 LIKE '5-011%' OR code1 LIKE '5-012%' OR code1 LIKE '5-013%' OR code1 LIKE '5-014%' OR code1 LIKE '5-015%' OR code1
LIKE '5-016%'
      OR code2 LIKE '5-011%' OR code2 LIKE '5-012%' OR code2 LIKE '5-013%' OR code2 LIKE '5-014%' OR code2 LIKE '5-015%' OR code2
LIKE '5-016%'
    )
```

```
  GROUP BY dx.patient_id
) q
```

**Reference SQL (Impala, HDP):**

```
WITH c71 AS (
  SELECT c_patnr, min(c_diagnose_timestamp) AS first_dx
  FROM db_tumpseudoprog_prepared.it_ishmed_diagnose
  WHERE c_diagnose_1 LIKE 'C71%' OR c_diagnose_2 LIKE 'C71%'
  GROUP BY c_patnr),
bd AS (SELECT DISTINCT c_patnr, c_birthdate FROM db_tumpseudoprog_prepared.it_ishmed_patient),
coh AS (
  SELECT c71.c_patnr, c71.first_dx,
         (year(c71.first_dx) - year(CAST(bd.c_birthdate AS TIMESTAMP))) AS age_dx
  FROM c71 JOIN bd ON c71.c_patnr = bd.c_patnr
  WHERE (year(c71.first_dx) - year(CAST(bd.c_birthdate AS TIMESTAMP))) >= 18),
res_events AS (
  SELECT c_patnr, CAST(c_prozedur_begin AS DATE) AS res_date
  FROM db_tumpseudoprog_prepared.it_ishmed_procedure
  WHERE regexp_like(c_prozedur_code,'^5-01[1-6]')
  GROUP BY c_patnr, CAST(c_prozedur_begin AS DATE))
SELECT count(*) AS n_re_resected FROM (
  SELECT re.c_patnr FROM res_events re JOIN coh ON re.c_patnr = coh.c_patnr
  GROUP BY re.c_patnr HAVING count(*) >= 2) t;
```

#### V5 — First diagnoses per year 2018-2023

*Outcome (Table 2): primary thread — incorrect and unlogged; query logged: no. Ratings DD / GG / consensus: 4 / 4 / 4. Re-test (new thread): not answered; query logged: —. Ratings DD / GG / consensus: 5 / 5 / 5.*

**Prompt(s) as posed:**

Among adult C71 patients, how many had their FIRST C71 diagnosis in each calendar year from 2018 to 2023? Give one count per year.

Platform result: 2018:286 2019:269 2020:255 2021:238 2022:276 2023:270 (sum 1594) | Impala reference: 2018:286 2019:247 2020:266 2021:238 2022:282 2023:268 (sum 1587)

**Generated SQL, primary thread: no query logged.**

Re-test (new thread, 1 Sep 2026): platform result could not complete

**Generated SQL, re-test: no query logged.**

**Reference SQL (Impala, HDP):**

```
WITH c71 AS (
  SELECT c_patnr, min(c_diagnose_timestamp) AS first_dx
  FROM db_tumpseudoprog_prepared.it_ishmed_diagnose
  WHERE c_diagnose_1 LIKE 'C71%' OR c_diagnose_2 LIKE 'C71%'
  GROUP BY c_patnr),
bd AS (SELECT DISTINCT c_patnr, c_birthdate FROM db_tumpseudoprog_prepared.it_ishmed_patient),
coh AS (
  SELECT c71.c_patnr, c71.first_dx,
         (year(c71.first_dx) - year(CAST(bd.c_birthdate AS TIMESTAMP))) AS age_dx
  FROM c71 JOIN bd ON c71.c_patnr = bd.c_patnr
  WHERE (year(c71.first_dx) - year(CAST(bd.c_birthdate AS TIMESTAMP))) >= 18)
SELECT year(first_dx) AS dx_year, count(*) AS n_patients
FROM coh
WHERE year(first_dx) BETWEEN 2018 AND 2023
GROUP BY year(first_dx) ORDER BY dx_year;
```

#### V6 — Top concomitant diagnoses (non-C71)

*Outcome (Table 2): primary thread — incorrect but traceable; query logged: yes. Ratings DD / GG / consensus: 2 / 3 / 3.*

**Prompt(s) as posed:**

Among adult C71 patients, what are the 10 most frequent OTHER diagnosis codes (excluding C71.x) recorded for these patients? Count distinct patients per code, order descending.

Platform result: Top: (nan 1594), G93.6 1347, Z11 1058, U99.0 817, D43.2 742, Z74.0 563, G40.8 482, U50.30 480, I10.90 464, U51.10 457 | Impala reference: G93.6 1347, Z11 1058, U99.0 817, D43.2 732, Z74.0 562, G40.8 476, U50.30 472, U51.10 457, I10.90 451, G93.5 432

**Generated SQL, primary thread — log entry Aug 25 2026 22:32 · status: data available · rows: 10**

```
SELECT diagnosis_code, COUNT(DISTINCT patient_id) AS distinct_patient_count
FROM (
    SELECT cohort.patient_id, dx.diagnosis_code
    FROM (
        SELECT adult.patient_id
        FROM (
```

```
            SELECT first_c71.patient_id, first_c71.first_c71_date, p.birthDate
            FROM (
                SELECT patient_id, MIN(recorded_dt) AS first_c71_date
                FROM (
                    SELECT
                        json_extract_scalar(c.subject, '$.identifier.value') AS patient_id,
                        json_extract_scalar(c.code, '$.coding[0].code') AS c0_code,
                        json_extract_scalar(c.code, '$.coding[1].code') AS c1_code,
                        json_extract_scalar(c.code, '$.coding[2].code') AS c2_code,
                        date_parse(substr(c.recordedDate, 1, 10), '%Y-%m-%d') AS recorded_dt
                    FROM condition c
                    WHERE c.recordedDate IS NOT NULL
                ) base
                WHERE recorded_dt IS NOT NULL
                  AND (
                        c0_code IN ('C71.0','C71.1','C71.2','C71.3','C71.4','C71.5','C71.6','C71.7','C71.8','C71.9')
                     OR c1_code IN ('C71.0','C71.1','C71.2','C71.3','C71.4','C71.5','C71.6','C71.7','C71.8','C71.9')
                     OR c2_code IN ('C71.0','C71.1','C71.2','C71.3','C71.4','C71.5','C71.6','C71.7','C71.8','C71.9')
                  )
                GROUP BY patient_id
            ) first_c71
            JOIN patient p
              ON json_extract_scalar(p.identifier, '$[0].value') = first_c71.patient_id
            WHERE p.birthDate IS NOT NULL
        ) adult
        WHERE date_diff('year', date_parse(substr(adult.birthDate, 1, 10), '%Y-%m-%d'), adult.first_c71_date) >= 18
    ) cohort
    JOIN (
        SELECT
            json_extract_scalar(c.subject, '$.identifier.value') AS patient_id,
            json_extract_scalar(c.code, '$.coding[0].code') AS diagnosis_code
        FROM condition c
        WHERE json_extract_scalar(c.code, '$.coding[0].code') IS NOT NULL
          AND json_extract_scalar(c.code, '$.coding[0].code') NOT LIKE 'C71%'
        UNION ALL
        SELECT
            json_extract_scalar(c.subject, '$.identifier.value') AS patient_id,
            json_extract_scalar(c.code, '$.coding[1].code') AS diagnosis_code
        FROM condition c
        WHERE json_extract_scalar(c.code, '$.coding[1].code') IS NOT NULL
          AND json_extract_scalar(c.code, '$.coding[1].code') NOT LIKE 'C71%'
        UNION ALL
        SELECT
            json_extract_scalar(c.subject, '$.identifier.value') AS patient_id,
            json_extract_scalar(c.code, '$.coding[2].code') AS diagnosis_code
        FROM condition c
        WHERE json_extract_scalar(c.code, '$.coding[2].code') IS NOT NULL
          AND json_extract_scalar(c.code, '$.coding[2].code') NOT LIKE 'C71%'
    ) dx
      ON cohort.patient_id = dx.patient_id
) counted
GROUP BY diagnosis_code
ORDER BY distinct_patient_count DESC, diagnosis_code
LIMIT 10
```

**Reference SQL (Impala, HDP):**

```
WITH c71 AS (
  SELECT c_patnr, min(c_diagnose_timestamp) AS first_dx
  FROM db_tumpseudoprog_prepared.it_ishmed_diagnose
  WHERE c_diagnose_1 LIKE 'C71%' OR c_diagnose_2 LIKE 'C71%'
  GROUP BY c_patnr),
bd AS (SELECT DISTINCT c_patnr, c_birthdate FROM db_tumpseudoprog_prepared.it_ishmed_patient),
coh AS (
  SELECT c71.c_patnr, c71.first_dx,
         (year(c71.first_dx) - year(CAST(bd.c_birthdate AS TIMESTAMP))) AS age_dx
  FROM c71 JOIN bd ON c71.c_patnr = bd.c_patnr
  WHERE (year(c71.first_dx) - year(CAST(bd.c_birthdate AS TIMESTAMP))) >= 18)
SELECT d.c_diagnose_1 AS icd, count(DISTINCT d.c_patnr) AS n_patients
FROM db_tumpseudoprog_prepared.it_ishmed_diagnose d JOIN coh ON d.c_patnr = coh.c_patnr
WHERE d.c_diagnose_1 IS NOT NULL AND d.c_diagnose_1 NOT LIKE 'C71%'
GROUP BY d.c_diagnose_1 ORDER BY n_patients DESC LIMIT 10;
```

#### V7 — Age distribution in decade bands

*Outcome (Table 2): primary thread — not answered; query logged: no. Ratings DD / GG / consensus: 5 / 4 / 5. Re-test (new thread): exact; query logged: yes. Ratings DD / GG / consensus: 1 / 1 / 1.*

**Prompt(s) as posed:**

Among adult patients (18 or older at first C71 diagnosis) with a glioblastoma diagnosis (ICD-10 C71.0 to C71.9 in any coding slot, regardless of verification status), how are patients distributed across age-at-first-diagnosis bands 18-29, 30-39, 40-49, 50-59, 60-69, 70-79, and 80 or older? Count distinct patients per band.

Platform result: distribution: FAILED 3x; single-band control 60-69 = 494 (INCORRECT)  |  Impala reference: 18-29:136 30-39:209 40-49:238 50-59:310 60-69:337 70-79:249 80+:115 (sum 1594)

**Generated SQL, primary thread: no query logged.**

Re-test (new thread, 1 Sep 2026): platform result 136/209/238/310/337/249/115 (exact)

**Generated SQL, re-test — log entry Sep 01 2026 14:15 · status: data available · rows: 7**

```
SELECT
  age_band,
  COUNT(DISTINCT patient_id) AS patient_count
FROM (
  SELECT
    patient_id,
    CASE
      WHEN age_years BETWEEN 18 AND 29 THEN '18-29'
      WHEN age_years BETWEEN 30 AND 39 THEN '30-39'
      WHEN age_years BETWEEN 40 AND 49 THEN '40-49'
      WHEN age_years BETWEEN 50 AND 59 THEN '50-59'
      WHEN age_years BETWEEN 60 AND 69 THEN '60-69'
      WHEN age_years BETWEEN 70 AND 79 THEN '70-79'
      WHEN age_years >= 80 THEN '80+'
      ELSE NULL
    END AS age_band
  FROM (
    SELECT
      patient_id,
      first_c71_date,
      date_diff(
        'year',
        date_parse(substr(birthDate, 1, 10), '%Y-%m-%d'),
        date_parse(substr(first_c71_date, 1, 10), '%Y-%m-%d')
      ) AS age_years
    FROM (
      SELECT
        dx.patient_id,
        dx.first_c71_date,
        p.birthDate
      FROM (
        SELECT
          patient_id,
          MIN(recordedDate) AS first_c71_date
        FROM (
          SELECT
            json_extract_scalar(c.subject, '$.identifier.value') AS patient_id,
            c.recordedDate,
            json_extract_scalar(c.code, '$.coding[0].code') AS code0,
            json_extract_scalar(c.code, '$.coding[1].code') AS code1,
            json_extract_scalar(c.code, '$.coding[2].code') AS code2
          FROM condition c
          WHERE
            json_extract_scalar(c.code, '$.coding[0].code') LIKE 'C71%'
            OR json_extract_scalar(c.code, '$.coding[1].code') LIKE 'C71%'
            OR json_extract_scalar(c.code, '$.coding[2].code') LIKE 'C71%'
        ) c71
        GROUP BY patient_id
      ) dx
      JOIN patient p
        ON json_extract_scalar(p.identifier, '$[0].value') = dx.patient_id
      WHERE dx.first_c71_date IS NOT NULL
        AND p.birthDate IS NOT NULL
    ) joined_data
  ) aged
  WHERE age_years >= 18
) binned
WHERE age_band IS NOT NULL
GROUP BY age_band
```

**Reference SQL (Impala, HDP):**

```
WITH c71 AS (
  SELECT c_patnr, min(c_diagnose_timestamp) AS first_dx
  FROM db_tumpseudoprog_prepared.it_ishmed_diagnose
  WHERE c_diagnose_1 LIKE 'C71%' OR c_diagnose_2 LIKE 'C71%'
  GROUP BY c_patnr),
bd AS (SELECT DISTINCT c_patnr, c_birthdate FROM db_tumpseudoprog_prepared.it_ishmed_patient),
coh AS (
```

```
  SELECT c71.c_patnr, c71.first_dx,
         (year(c71.first_dx) - year(CAST(bd.c_birthdate AS TIMESTAMP))) AS age_dx
  FROM c71 JOIN bd ON c71.c_patnr = bd.c_patnr
  WHERE (year(c71.first_dx) - year(CAST(bd.c_birthdate AS TIMESTAMP))) >= 18)
SELECT CASE WHEN age_dx BETWEEN 18 AND 29 THEN '18-29' WHEN age_dx BETWEEN 30 AND 39 THEN '30-39'
  WHEN age_dx BETWEEN 40 AND 49 THEN '40-49' WHEN age_dx BETWEEN 50 AND 59 THEN '50-59'
  WHEN age_dx BETWEEN 60 AND 69 THEN '60-69' WHEN age_dx BETWEEN 70 AND 79 THEN '70-79'
  ELSE '80+' END AS band, count(*) AS n
FROM coh GROUP BY 1 ORDER BY 1;
```

#### V8 — Sex x age band (2-way grouping)

*Outcome (Table 2): primary thread — exact; query logged: yes. Ratings DD / GG / consensus: 1 / 1 / 1.*

**Prompt(s) as posed:**

For the same adult C71 cohort, break down the number of distinct patients by sex AND age band (18-49, 50-64, 65+) at first diagnosis. One count per sex-band combination.

Platform result: 1st: sum 113 (silent filter); 2nd: F 249/206/217, M 334/288/300 (sum 1594) | Impala reference: F 249/206/217, M 334/288/300 (sum 1594)

**Generated SQL, primary thread — log entry Sep 01 2026 12:45 · status: data available · rows: 6**

```
SELECT
  sex,
  age_band,
  COUNT(DISTINCT patient_id) AS patient_count
FROM (
  SELECT
    d.patient_id,
    p.gender AS sex,
    CASE
      WHEN date_diff('year', date_parse(substr(p.birthDate, 1, 10), '%Y-%m-%d'), date_parse(substr(d.first_dx_date, 1, 10), '%Y-
%m-%d')) BETWEEN 18 AND 49 THEN '18-49'
      WHEN date_diff('year', date_parse(substr(p.birthDate, 1, 10), '%Y-%m-%d'), date_parse(substr(d.first_dx_date, 1, 10), '%Y-
%m-%d')) BETWEEN 50 AND 64 THEN '50-64'
      WHEN date_diff('year', date_parse(substr(p.birthDate, 1, 10), '%Y-%m-%d'), date_parse(substr(d.first_dx_date, 1, 10), '%Y-
%m-%d')) >= 65 THEN '65+'
      ELSE NULL
    END AS age_band
  FROM (
    SELECT
      patient_id,
      MIN(recorded_date) AS first_dx_date
    FROM (
      SELECT
        json_extract_scalar(c.subject, '$.identifier.value') AS patient_id,
        c.recordedDate AS recorded_date
      FROM condition c
      WHERE json_extract_scalar(c.verificationStatus, '$.text') = 'G'
        AND (
          json_extract_scalar(c.code, '$.coding[0].code') LIKE 'C71%'
          OR json_extract_scalar(c.code, '$.coding[1].code') LIKE 'C71%'
          OR json_extract_scalar(c.code, '$.coding[2].code') LIKE 'C71%'
        )
    ) q
    GROUP BY patient_id
  ) d
  JOIN patient p
    ON json_extract_scalar(p.identifier, '$[0].value') = d.patient_id
  WHERE p.gender IN ('F', 'M')
    AND p.birthDate IS NOT NULL
    AND date_diff('year', date_parse(substr(p.birthDate, 1, 10), '%Y-%m-%d'), date_parse(substr(d.first_dx_date, 1, 10), '%Y-%m-
%d')) >= 18
) x
WHERE age_band IS NOT NULL
GROUP BY sex, age_band
ORDER BY sex, age_band
```

**Generated SQL, primary thread — log entry Sep 01 2026 12:50 · status: data available · rows: 6**

```
SELECT
  sex,
  age_band,
  COUNT(DISTINCT patient_id) AS patient_count
FROM (
  SELECT
    patient_id,
    sex,
    age_at_first_dx,
```

```
    CASE
      WHEN age_at_first_dx BETWEEN 18 AND 49 THEN '18-49'
      WHEN age_at_first_dx BETWEEN 50 AND 64 THEN '50-64'
      WHEN age_at_first_dx >= 65 THEN '65+'
      ELSE NULL
    END AS age_band
  FROM (
    SELECT
      patient_id,
      sex,
      date_diff('year', birth_dt, first_dx_dt) AS age_at_first_dx
    FROM (
      SELECT
        patient_id,
        sex,
        birth_dt,
        first_dx_dt
      FROM (
        SELECT
          patient_id,
          sex,
          birth_dt,
          first_dx_dt,
          ROW_NUMBER() OVER (PARTITION BY patient_id ORDER BY first_dx_dt ASC) AS rn
        FROM (
          SELECT
            json_extract_scalar(c.subject, '$.identifier.value') AS patient_id,
            p.gender AS sex,
            date_parse(substr(p.birthDate, 1, 10), '%Y-%m-%d') AS birth_dt,
            date_parse(substr(c.recordedDate, 1, 10), '%Y-%m-%d') AS first_dx_dt
          FROM condition c
          JOIN patient p
            ON json_extract_scalar(c.subject, '$.identifier.value') = json_extract_scalar(p.identifier, '$[0].value')
          WHERE (
                 json_extract_scalar(c.code, '$.coding[0].code') IN
('C71.0','C71.1','C71.2','C71.3','C71.4','C71.5','C71.6','C71.7','C71.8','C71.9')
              OR json_extract_scalar(c.code, '$.coding[1].code') IN
('C71.0','C71.1','C71.2','C71.3','C71.4','C71.5','C71.6','C71.7','C71.8','C71.9')
              OR json_extract_scalar(c.code, '$.coding[2].code') IN
('C71.0','C71.1','C71.2','C71.3','C71.4','C71.5','C71.6','C71.7','C71.8','C71.9')
          )
            AND p.birthDate IS NOT NULL
            AND c.recordedDate IS NOT NULL
        ) extracted
      ) ranked
      WHERE rn = 1
    ) first_dx
    WHERE date_diff('year', birth_dt, first_dx_dt) >= 18
  ) aged
) banded
WHERE age_band IS NOT NULL
GROUP BY sex, age_band
ORDER BY sex, age_band
```

**Reference SQL (Impala, HDP):**

```
WITH c71 AS (
  SELECT c_patnr, min(c_diagnose_timestamp) AS first_dx
  FROM db_tumpseudoprog_prepared.it_ishmed_diagnose
  WHERE c_diagnose_1 LIKE 'C71%' OR c_diagnose_2 LIKE 'C71%'
  GROUP BY c_patnr),
bd AS (SELECT DISTINCT c_patnr, c_birthdate FROM db_tumpseudoprog_prepared.it_ishmed_patient),
coh AS (
  SELECT c71.c_patnr, c71.first_dx,
         (year(c71.first_dx) - year(CAST(bd.c_birthdate AS TIMESTAMP))) AS age_dx
  FROM c71 JOIN bd ON c71.c_patnr = bd.c_patnr
  WHERE (year(c71.first_dx) - year(CAST(bd.c_birthdate AS TIMESTAMP))) >= 18),
g AS (SELECT DISTINCT c_patnr, c_gender FROM db_tumpseudoprog_prepared.it_ishmed_patient)
SELECT g.c_gender, CASE WHEN coh.age_dx<50 THEN '18-49' WHEN coh.age_dx<65 THEN '50-64' ELSE '65+' END AS band,
       count(DISTINCT coh.c_patnr) AS n
FROM coh JOIN g ON coh.c_patnr=g.c_patnr
GROUP BY 1,2 ORDER BY 1,2;
```

#### V9 — Hospital cases per patient (median)

*Outcome (Table 2): primary thread — exact; query logged: yes. Ratings DD / GG / consensus: 1 / 1 / 1.*

**Prompt(s) as posed:**

For the adult C71 cohort, how many distinct hospital cases (encounters) does each patient have, and what is the median number of cases per patient?

Platform result: n=1594; median 1; IQR 1-2; min 1; max 5 | Impala reference: n=1594; median 1; IQR 1-1; min 1; max 5; mean 1.21 (83% single case)

**Generated SQL, primary thread — log entry Sep 01 2026 13:46 · status: data available · rows: 1936**

```
SELECT
  q.patient_id,
  q.first_c71_date,
  COALESCE(ec.encounter_count, 0) AS encounter_count
FROM (
  SELECT
    dx.patient_id,
    MIN(dx.recorded_date) AS first_c71_date
  FROM (
    SELECT
      json_extract_scalar(c.subject, '$.identifier.value') AS patient_id,
      substr(c.recordedDate, 1, 10) AS recorded_date
    FROM condition c
    WHERE
      json_extract_scalar(c.code, '$.coding[0].code') IN
('C71.0','C71.1','C71.2','C71.3','C71.4','C71.5','C71.6','C71.7','C71.8','C71.9')
      OR json_extract_scalar(c.code, '$.coding[1].code') IN
('C71.0','C71.1','C71.2','C71.3','C71.4','C71.5','C71.6','C71.7','C71.8','C71.9')
      OR json_extract_scalar(c.code, '$.coding[2].code') IN
('C71.0','C71.1','C71.2','C71.3','C71.4','C71.5','C71.6','C71.7','C71.8','C71.9')
  ) dx
  GROUP BY dx.patient_id
) q
JOIN patient p
  ON json_extract_scalar(p.identifier, '$[0].value') = q.patient_id
LEFT JOIN (
  SELECT
    json_extract_scalar(e.subject, '$.identifier.value') AS patient_id,
    COUNT(DISTINCT e.id) AS encounter_count
  FROM encounter e
  GROUP BY json_extract_scalar(e.subject, '$.identifier.value')
) ec
  ON ec.patient_id = q.patient_id
WHERE date_diff(
        'year',
        date_parse(substr(p.birthDate, 1, 10), '%Y-%m-%d'),
        date_parse(q.first_c71_date, '%Y-%m-%d')
      ) >= 18
```

**Reference SQL (Impala, HDP):**

```
WITH c71 AS (
  SELECT c_patnr, min(c_diagnose_timestamp) AS first_dx
  FROM db_tumpseudoprog_prepared.it_ishmed_diagnose
  WHERE c_diagnose_1 LIKE 'C71%' OR c_diagnose_2 LIKE 'C71%'
  GROUP BY c_patnr),
bd AS (SELECT DISTINCT c_patnr, c_birthdate FROM db_tumpseudoprog_prepared.it_ishmed_patient),
coh AS (
  SELECT c71.c_patnr
  FROM c71 JOIN bd ON c71.c_patnr = bd.c_patnr
  WHERE (year(c71.first_dx) - year(CAST(bd.c_birthdate AS TIMESTAMP))) >= 18)
SELECT coh.c_patnr, count(DISTINCT d.c_falnr) AS n_cases
FROM coh JOIN db_tumpseudoprog_prepared.it_ishmed_diagnose d ON coh.c_patnr = d.c_patnr
WHERE d.c_falnr IS NOT NULL
GROUP BY coh.c_patnr;
-- EXPORT rows; median offline. Cases = distinct c_falnr in DIAGNOSIS table (all coded cases; confirmed via DESCRIBE)
```

#### V10 — Distinct diagnosis codes per patient (comorbidity load)

*Outcome (Table 2): primary thread — not answered; query logged: —. Ratings DD / GG / consensus: 5 / 5 / 5. Re-test (new thread): concordant after definition alignment; query logged: yes. Ratings DD / GG / consensus: 2 / 3 / 2.*

**Prompt(s) as posed:**

For the adult C71 cohort, how many distinct diagnosis codes does each patient have, and what is the median per patient?

Platform result: FAILED 2x: (1) explained abstention - variable not available; (2) frequency-table reprompt -> couldn't complete | Impala reference: n=1594; median 12; IQR 8-17; min 2; max 91; mean 13.8; 59% >10 codes

**Generated SQL, primary thread: no query logged.**

Re-test (new thread, 1 Sep 2026): platform result n=1,594; median 13; IQR 9–18; min 3; max 92

##### Generated SQL, re-test — log entry Sep 01 2026 14:43 • status: data available • rows: 1594

```
SELECT
  patient_id,
  first_c71_date,
  birthDate,
  COUNT(DISTINCT dx_code) AS unique_diagnosis_code_count
FROM (
  SELECT
    codes.patient_id,
    firsts.first_c71_date,
    p.birthDate,
    codes.dx_code
  FROM (
    SELECT
      patient_id,
      MIN(recorded_dt) AS first_c71_date
    FROM (
      SELECT
        json_extract_scalar(c.subject, '$.identifier.value') AS patient_id,
        date_parse(substr(c.recordedDate, 1, 10), '%Y-%m-%d') AS recorded_dt
      FROM condition c
      WHERE
        json_extract_scalar(c.code, '$.coding[0].code') IN
('C71.0','C71.1','C71.2','C71.3','C71.4','C71.5','C71.6','C71.7','C71.8','C71.9')
        OR json_extract_scalar(c.code, '$.coding[1].code') IN
('C71.0','C71.1','C71.2','C71.3','C71.4','C71.5','C71.6','C71.7','C71.8','C71.9')
        OR json_extract_scalar(c.code, '$.coding[2].code') IN
('C71.0','C71.1','C71.2','C71.3','C71.4','C71.5','C71.6','C71.7','C71.8','C71.9')
    ) c71
    GROUP BY patient_id
  ) firsts
  JOIN patient p
    ON json_extract_scalar(p.identifier, '$[0].value') = firsts.patient_id
  JOIN (
    SELECT
      json_extract_scalar(c.subject, '$.identifier.value') AS patient_id,
      json_extract_scalar(c.code, '$.coding[0].code') AS dx_code
    FROM condition c
    WHERE json_extract_scalar(c.code, '$.coding[0].code') IS NOT NULL

    UNION ALL

    SELECT
      json_extract_scalar(c.subject, '$.identifier.value') AS patient_id,
      json_extract_scalar(c.code, '$.coding[1].code') AS dx_code
    FROM condition c
    WHERE json_extract_scalar(c.code, '$.coding[1].code') IS NOT NULL

    UNION ALL

    SELECT
      json_extract_scalar(c.subject, '$.identifier.value') AS patient_id,
      json_extract_scalar(c.code, '$.coding[2].code') AS dx_code
    FROM condition c
    WHERE json_extract_scalar(c.code, '$.coding[2].code') IS NOT NULL
  ) codes
    ON codes.patient_id = firsts.patient_id
  WHERE date_diff('year', date_parse(substr(p.birthDate, 1, 10), '%Y-%m-%d'), firsts.first_c71_date) >= 18
) t
GROUP BY patient_id, first_c71_date, birthDate
```

##### Generated SQL, re-test — log entry Sep 01 2026 14:44 • status: data available • rows: 1594

```
SELECT
  patient_id,
  first_c71_date,
  unique_diagnosis_code_count
FROM (
  SELECT
    firsts.patient_id,
    firsts.first_c71_date,
    COUNT(DISTINCT codes.dx_code) AS unique_diagnosis_code_count
  FROM (
    SELECT
      c71.patient_id,
      MIN(c71.recorded_dt) AS first_c71_date
    FROM (
      SELECT
        json_extract_scalar(c.subject, '$.identifier.value') AS patient_id,
        date_parse(substr(c.recordedDate, 1, 10), '%Y-%m-%d') AS recorded_dt
      FROM condition c
```

```
      WHERE
        json_extract_scalar(c.code, '$.coding[0].code') IN
('C71.0','C71.1','C71.2','C71.3','C71.4','C71.5','C71.6','C71.7','C71.8','C71.9')
        OR json_extract_scalar(c.code, '$.coding[1].code') IN
('C71.0','C71.1','C71.2','C71.3','C71.4','C71.5','C71.6','C71.7','C71.8','C71.9')
        OR json_extract_scalar(c.code, '$.coding[2].code') IN
('C71.0','C71.1','C71.2','C71.3','C71.4','C71.5','C71.6','C71.7','C71.8','C71.9')
    ) c71
    GROUP BY c71.patient_id
  ) firsts
  JOIN patient p
    ON json_extract_scalar(p.identifier, '$[0].value') = firsts.patient_id
  JOIN (
    SELECT
      json_extract_scalar(c.subject, '$.identifier.value') AS patient_id,
      json_extract_scalar(c.code, '$.coding[0].code') AS dx_code
    FROM condition c
    WHERE json_extract_scalar(c.code, '$.coding[0].code') IS NOT NULL

    UNION ALL

    SELECT
      json_extract_scalar(c.subject, '$.identifier.value') AS patient_id,
      json_extract_scalar(c.code, '$.coding[1].code') AS dx_code
    FROM condition c
    WHERE json_extract_scalar(c.code, '$.coding[1].code') IS NOT NULL

    UNION ALL

    SELECT
      json_extract_scalar(c.subject, '$.identifier.value') AS patient_id,
      json_extract_scalar(c.code, '$.coding[2].code') AS dx_code
    FROM condition c
    WHERE json_extract_scalar(c.code, '$.coding[2].code') IS NOT NULL
  ) codes
    ON codes.patient_id = firsts.patient_id
  WHERE date_diff('year', date_parse(substr(p.birthDate, 1, 10), '%Y-%m-%d'), firsts.first_c71_date) >= 18
  GROUP BY firsts.patient_id, firsts.first_c71_date
) t
```

**Reference SQL (Impala, HDP):**

```
WITH c71 AS (
  SELECT c_patnr, min(c_diagnose_timestamp) AS first_dx
  FROM db_tumpseudoprog_prepared.it_ishmed_diagnose
  WHERE c_diagnose_1 LIKE 'C71%' OR c_diagnose_2 LIKE 'C71%'
  GROUP BY c_patnr),
bd AS (SELECT DISTINCT c_patnr, c_birthdate FROM db_tumpseudoprog_prepared.it_ishmed_patient),
coh AS (
  SELECT c71.c_patnr, c71.first_dx,
         (year(c71.first_dx) - year(CAST(bd.c_birthdate AS TIMESTAMP))) AS age_dx
  FROM c71 JOIN bd ON c71.c_patnr = bd.c_patnr
  WHERE (year(c71.first_dx) - year(CAST(bd.c_birthdate AS TIMESTAMP))) >= 18)
SELECT coh.c_patnr, count(DISTINCT d.c_diagnose_1) AS n_codes
FROM coh JOIN db_tumpseudoprog_prepared.it_ishmed_diagnose d ON coh.c_patnr=d.c_patnr
WHERE d.c_diagnose_1 IS NOT NULL
GROUP BY coh.c_patnr;
-- EXPORT rows; median offline
```

**Reference SQL, second variant (platform definition; basis for the re-test classification 2):**

```
-- v1.2, variant counting distinct codes across all three coding slots mapped to Condition.code.coding[0..2]
-- (c_diagnose_1, c_diagnose_2, c_orphacode). Reproduces the platform result of the re-test exactly: n 1594, median 13, min 3, max
92 (Impala, 16 Oct 2026).
WITH c71 AS (
  SELECT c_patnr, min(c_diagnose_timestamp) AS first_dx
  FROM db_tumpseudoprog_prepared.it_ishmed_diagnose
  WHERE c_diagnose_1 LIKE 'C71%' OR c_diagnose_2 LIKE 'C71%'
  GROUP BY c_patnr),
bd AS (SELECT DISTINCT c_patnr, c_birthdate FROM db_tumpseudoprog_prepared.it_ishmed_patient),
coh AS (
  SELECT c71.c_patnr
  FROM c71 JOIN bd ON c71.c_patnr = bd.c_patnr
  WHERE (year(c71.first_dx) - year(CAST(bd.c_birthdate AS TIMESTAMP))) >= 18),
codes AS (
  SELECT c_patnr, c_diagnose_1 AS code FROM db_tumpseudoprog_prepared.it_ishmed_diagnose WHERE c_diagnose_1 IS NOT NULL
  UNION ALL
  SELECT c_patnr, c_diagnose_2 FROM db_tumpseudoprog_prepared.it_ishmed_diagnose WHERE c_diagnose_2 IS NOT NULL
  UNION ALL
  SELECT c_patnr, c_orphacode FROM db_tumpseudoprog_prepared.it_ishmed_diagnose WHERE c_orphacode IS NOT NULL),
per_pat AS (
  SELECT coh.c_patnr, count(DISTINCT codes.code) AS n_codes
  FROM coh JOIN codes ON coh.c_patnr = codes.c_patnr
```

```
  GROUP BY coh.c_patnr)
SELECT count(*) AS n, appx_median(n_codes) AS median_codes, min(n_codes) AS min_codes, max(n_codes) AS max_codes
FROM per_pat;
```

#### V11 — MRI examinations per patient (median)

*Outcome (Table 2): primary thread — not answered; query logged: —. Ratings DD / GG / consensus: 5 / 5 / 5. Re-test (new thread): not answered; query logged: —. Ratings DD / GG / consensus: 5 / 5 / 5.*

**Prompt(s) as posed:**

For the adult C71 cohort, how many cranial MRI procedures (OPS code starting with 3-820) does each patient have on distinct dates, and what is the median per patient?

Platform result: FAILED: couldn't complete (count + distribution prompt) | Impala reference: n=1415 with >=1 MRI (=B4 top-1); median 2; IQR 1-2; max 9; 82% have 1-2

**Generated SQL, primary thread: no query logged.**

Re-test (new thread, 1 Sep 2026): platform result could not complete

**Generated SQL, re-test: no query logged.**

**Reference SQL (Impala, HDP):**

```
WITH c71 AS (
  SELECT c_patnr, min(c_diagnose_timestamp) AS first_dx
  FROM db_tumpseudoprog_prepared.it_ishmed_diagnose
  WHERE c_diagnose_1 LIKE 'C71%' OR c_diagnose_2 LIKE 'C71%'
  GROUP BY c_patnr),
bd AS (SELECT DISTINCT c_patnr, c_birthdate FROM db_tumpseudoprog_prepared.it_ishmed_patient),
coh AS (
  SELECT c71.c_patnr, c71.first_dx,
         (year(c71.first_dx) - year(CAST(bd.c_birthdate AS TIMESTAMP))) AS age_dx
  FROM c71 JOIN bd ON c71.c_patnr = bd.c_patnr
  WHERE (year(c71.first_dx) - year(CAST(bd.c_birthdate AS TIMESTAMP))) >= 18)
SELECT coh.c_patnr, count(DISTINCT CAST(pr.c_prozedur_begin AS DATE)) AS n_mri
FROM coh JOIN db_tumpseudoprog_prepared.it_ishmed_procedure pr ON coh.c_patnr=pr.c_patnr
WHERE pr.c_prozedur_code LIKE '3-820%'
GROUP BY coh.c_patnr;
-- EXPORT rows; median offline
```

#### V12 — Chemotherapy (OPS 8-54x): patients and share

*Outcome (Table 2): primary thread — incorrect and unlogged; query logged: no. Ratings DD / GG / consensus: 4 / 4 / 4. Re-test (new thread): not answered; query logged: —. Ratings DD / GG / consensus: 5 / 5 / 5.*

**Prompt(s) as posed:**

Among the adult C71 cohort, how many distinct patients have any chemotherapy procedure code (OPS starting with 8-54), and what share of the cohort is that?

Platform result: 512 / 1594 (32.1%) | Impala reference: 142 / 1594 (8.9%)

**Generated SQL, primary thread: no query logged.**

Re-test (new thread, 1 Sep 2026): platform result could not complete

**Generated SQL, re-test: no query logged.**

**Reference SQL (Impala, HDP):**

```
WITH c71 AS (
  SELECT c_patnr, min(c_diagnose_timestamp) AS first_dx
  FROM db_tumpseudoprog_prepared.it_ishmed_diagnose
  WHERE c_diagnose_1 LIKE 'C71%' OR c_diagnose_2 LIKE 'C71%'
  GROUP BY c_patnr),
bd AS (SELECT DISTINCT c_patnr, c_birthdate FROM db_tumpseudoprog_prepared.it_ishmed_patient),
coh AS (
  SELECT c71.c_patnr, c71.first_dx,
         (year(c71.first_dx) - year(CAST(bd.c_birthdate AS TIMESTAMP))) AS age_dx
  FROM c71 JOIN bd ON c71.c_patnr = bd.c_patnr
  WHERE (year(c71.first_dx) - year(CAST(bd.c_birthdate AS TIMESTAMP))) >= 18),
chemo AS (SELECT DISTINCT c_patnr FROM db_tumpseudoprog_prepared.it_ishmed_procedure WHERE c_prozedur_code LIKE '8-54%')
SELECT count(DISTINCT CASE WHEN ch.c_patnr IS NOT NULL THEN coh.c_patnr END) AS n_chemo,
       count(DISTINCT coh.c_patnr) AS n_cohort,
       round(100.0*count(DISTINCT CASE WHEN ch.c_patnr IS NOT NULL THEN coh.c_patnr END)/count(DISTINCT coh.c_patnr),1) AS pct
FROM coh LEFT JOIN chemo ch ON coh.c_patnr=ch.c_patnr;
```

#### V13 — Radiotherapy (OPS 8-52x): patients and share

*Outcome (Table 2): primary thread — incorrect and unlogged; query logged: no. Ratings DD / GG / consensus: 4 / 4 / 4. Re-test (new thread): exact; query logged: yes. Ratings DD / GG / consensus: 1 / 1 / 1.*

**Prompt(s) as posed:**

Among the adult C71 cohort, how many distinct patients have any radiotherapy procedure code (OPS starting with 8-52), and what share of the cohort is that?

Platform result: 684 / 1594 (42.9%) | Impala reference: 116 / 1594 (7.3%)

**Generated SQL, primary thread: no query logged.**

Re-test (new thread, 1 Sep 2026): platform result 116 / 1,594 (7.28%) exact

**Generated SQL, re-test — log entry Sep 01 2026 14:19 · status: data available · rows: 1594**

```
SELECT
  cohort.patient_id,
  CASE
    WHEN proc.patient_id IS NOT NULL THEN 1
    ELSE 0
  END AS has_ops_852
FROM (
  SELECT adult.patient_id
  FROM (
    SELECT
      first_c71.patient_id,
      date_diff(
        'year',
        date_parse(substr(first_c71.birthDate, 1, 10), '%Y-%m-%d'),
        date_parse(substr(first_c71.first_c71_date, 1, 10), '%Y-%m-%d')
      ) AS age_at_first_c71
    FROM (
      SELECT
        ranked.patient_id,
        ranked.birthDate,
        ranked.recordedDate AS first_c71_date
      FROM (
        SELECT
          extracted.patient_id,
          extracted.birthDate,
          extracted.recordedDate,
          ROW_NUMBER() OVER (
            PARTITION BY extracted.patient_id
            ORDER BY extracted.recordedDate ASC
          ) AS rn
        FROM (
          SELECT
            json_extract_scalar(c.subject, '$.identifier.value') AS patient_id,
            p.birthDate AS birthDate,
            c.recordedDate AS recordedDate
          FROM condition c
          JOIN patient p
            ON json_extract_scalar(c.subject, '$.identifier.value') = json_extract_scalar(p.identifier, '$[0].value')
          WHERE
            json_extract_scalar(c.code, '$.coding[0].code') LIKE 'C71%'
            OR json_extract_scalar(c.code, '$.coding[1].code') LIKE 'C71%'
            OR json_extract_scalar(c.code, '$.coding[2].code') LIKE 'C71%'
        ) extracted
      ) ranked
      WHERE ranked.rn = 1
    ) first_c71
  ) adult
  WHERE adult.age_at_first_c71 >= 18
) cohort
LEFT JOIN (
  SELECT DISTINCT
    json_extract_scalar(pr.subject, '$.identifier.value') AS patient_id
  FROM procedure pr
  WHERE
    json_extract_scalar(pr.code, '$.coding[0].code') LIKE '8-52%'
    OR json_extract_scalar(pr.code, '$.coding[1].code') LIKE '8-52%'
    OR json_extract_scalar(pr.code, '$.coding[2].code') LIKE '8-52%'
) proc
  ON cohort.patient_id = proc.patient_id
```

**Reference SQL (Impala, HDP):**

```
WITH c71 AS (
  SELECT c_patnr, min(c_diagnose_timestamp) AS first_dx
```

```
  FROM db_tumpseudoprog_prepared.it_ishmed_diagnose
  WHERE c_diagnose_1 LIKE 'C71%' OR c_diagnose_2 LIKE 'C71%'
  GROUP BY c_patnr),
bd AS (SELECT DISTINCT c_patnr, c_birthdate FROM db_tumpseudoprog_prepared.it_ishmed_patient),
coh AS (
  SELECT c71.c_patnr, c71.first_dx,
         (year(c71.first_dx) - year(CAST(bd.c_birthdate AS TIMESTAMP))) AS age_dx
  FROM c71 JOIN bd ON c71.c_patnr = bd.c_patnr
  WHERE (year(c71.first_dx) - year(CAST(bd.c_birthdate AS TIMESTAMP))) >= 18),
rt AS (SELECT DISTINCT c_patnr FROM db_tumpseudoprog_prepared.it_ishmed_procedure WHERE c_prozedur_code LIKE '8-52%')
SELECT count(DISTINCT CASE WHEN r.c_patnr IS NOT NULL THEN coh.c_patnr END) AS n_rt,
       count(DISTINCT coh.c_patnr) AS n_cohort,
       round(100.0*count(DISTINCT CASE WHEN r.c_patnr IS NOT NULL THEN coh.c_patnr END)/count(DISTINCT coh.c_patnr),1) AS pct
FROM coh LEFT JOIN rt r ON coh.c_patnr=r.c_patnr;
```

#### V14 — Hydrocephalus/shunt procedures (5-023x)

*Outcome (Table 2): primary thread — incorrect and unlogged; query logged: no. Ratings DD / GG / consensus: 4 / 4 / 4. Re-test (new thread): exact; query logged: yes. Ratings DD / GG / consensus: 1 / 1 / 1.*

**Prompt(s) as posed:**

Among the adult C71 cohort, how many distinct patients have a ventricular shunt or hydrocephalus-related procedure (OPS starting with 5-023)?

Platform result: 88 | Impala reference: 76

**Generated SQL, primary thread: no query logged.**

Re-test (new thread, 1 Sep 2026): platform result 76 / 1,594 (4.77%) exact

**Generated SQL, re-test — log entry Sep 01 2026 14:26 · status: data available · rows: 1**

```
SELECT
  COUNT(DISTINCT CASE WHEN proc.patient_id IS NOT NULL THEN adult.patient_id END) AS numerator,
  COUNT(DISTINCT adult.patient_id) AS denominator
FROM (
  SELECT patient_id
  FROM (
    SELECT
      dx.patient_id,
      dx.first_c71_date,
      date_diff(
        'year',
        date_parse(substr(p.birthDate, 1, 10), '%Y-%m-%d'),
        date_parse(substr(dx.first_c71_date, 1, 10), '%Y-%m-%d')
      ) AS age_at_first_c71
    FROM (
      SELECT
        patient_id,
        MIN(recorded_date) AS first_c71_date
      FROM (
        SELECT
          json_extract_scalar(c.subject, '$.identifier.value') AS patient_id,
          c.recordedDate AS recorded_date
        FROM condition c
        WHERE
          json_extract_scalar(c.code, '$.coding[0].code') IN
('C71.0','C71.1','C71.2','C71.3','C71.4','C71.5','C71.6','C71.7','C71.8','C71.9')
          OR json_extract_scalar(c.code, '$.coding[1].code') IN
('C71.0','C71.1','C71.2','C71.3','C71.4','C71.5','C71.6','C71.7','C71.8','C71.9')
          OR json_extract_scalar(c.code, '$.coding[2].code') IN
('C71.0','C71.1','C71.2','C71.3','C71.4','C71.5','C71.6','C71.7','C71.8','C71.9')
      ) c71
      WHERE recorded_date IS NOT NULL
      GROUP BY patient_id
    ) dx
    JOIN patient p
      ON json_extract_scalar(p.identifier, '$[0].value') = dx.patient_id
    WHERE p.birthDate IS NOT NULL
  ) aged
  WHERE age_at_first_c71 >= 18
) adult
LEFT JOIN (
  SELECT DISTINCT
    json_extract_scalar(pr.subject, '$.identifier.value') AS patient_id
  FROM procedure pr
  WHERE
    json_extract_scalar(pr.code, '$.coding[0].code') LIKE '5-023%'
    OR json_extract_scalar(pr.code, '$.coding[1].code') LIKE '5-023%'
    OR json_extract_scalar(pr.code, '$.coding[2].code') LIKE '5-023%'
```

```
) proc
  ON adult.patient_id = proc.patient_id
```

**Reference SQL (Impala, HDP):**

```
WITH c71 AS (
  SELECT c_patnr, min(c_diagnose_timestamp) AS first_dx
  FROM db_tumpseudoprog_prepared.it_ishmed_diagnose
  WHERE c_diagnose_1 LIKE 'C71%' OR c_diagnose_2 LIKE 'C71%'
  GROUP BY c_patnr),
bd AS (SELECT DISTINCT c_patnr, c_birthdate FROM db_tumpseudoprog_prepared.it_ishmed_patient),
coh AS (
  SELECT c71.c_patnr, c71.first_dx,
         (year(c71.first_dx) - year(CAST(bd.c_birthdate AS TIMESTAMP))) AS age_dx
  FROM c71 JOIN bd ON c71.c_patnr = bd.c_patnr
  WHERE (year(c71.first_dx) - year(CAST(bd.c_birthdate AS TIMESTAMP))) >= 18),
sh AS (SELECT DISTINCT c_patnr FROM db_tumpseudoprog_prepared.it_ishmed_procedure WHERE c_prozedur_code LIKE '5-023%')
SELECT count(DISTINCT coh.c_patnr) AS n_shunt
FROM coh JOIN sh ON coh.c_patnr=sh.c_patnr;
```

#### V15 — Days first diagnosis to death (deceased only)

*Outcome (Table 2): primary thread — incorrect and unlogged; query logged: no. Ratings DD / GG / consensus: 4 / 4 / 4. Re-test (new thread): incorrect but traceable; query logged: yes (display ≠ query). Ratings DD / GG / consensus: 3 / 2 / 3.*

**Prompt(s) as posed:**

Among the adult C71 patients who are deceased, what is the median number of days from first C71 diagnosis to death? State how many deceased patients are included.

Platform result: n=101; median 231 d; IQR 74-517 | Impala reference: n=101; median 43 d; IQR 10-275; mean 220; 44% die within 30 d

**Generated SQL, primary thread: no query logged.**

Re-test (new thread, 1 Sep 2026): platform result n=84; median 93 d; IQR 20.2–339.5

*[Re-test classification: the logged query returned 115 patient-level rows (including duplicates) for 101 distinct patients, but statistics were displayed for 84 patients; the reduction from 101 to 84 has no counterpart in the query. Classified as incorrect but traceable and marked separately in Table 2 (display ≠ query).]*

**Generated SQL, re-test — log entry Sep 01 2026 14:41 · status: data available · rows: 115**

```
SELECT
  q.patient_id,
  q.first_dx_date,
  q.deceased_dt,
  q.birth_date
FROM (
  SELECT
    patient_id,
    first_dx_date,
    deceased_dt,
    birth_date,
    date_diff('year', date_parse(substr(birth_date, 1, 10), '%Y-%m-%d'), date_parse(substr(first_dx_date, 1, 10), '%Y-%m-%d')) AS
age_at_dx
  FROM (
    SELECT
      json_extract_scalar(p.identifier, '$[0].value') AS patient_id,
      MIN(c.recordedDate) AS first_dx_date,
      p.deceasedDateTime AS deceased_dt,
      p.birthDate AS birth_date
    FROM patient p
    JOIN condition c
      ON json_extract_scalar(c.subject, '$.identifier.value') = json_extract_scalar(p.identifier, '$[0].value')
    WHERE p.deceasedDateTime IS NOT NULL
      AND p.birthDate IS NOT NULL
      AND c.recordedDate IS NOT NULL
      AND (
        json_extract_scalar(c.code, '$.coding[0].code') IN
('C71.0','C71.1','C71.2','C71.3','C71.4','C71.5','C71.6','C71.7','C71.8','C71.9')
        OR json_extract_scalar(c.code, '$.coding[1].code') IN
('C71.0','C71.1','C71.2','C71.3','C71.4','C71.5','C71.6','C71.7','C71.8','C71.9')
        OR json_extract_scalar(c.code, '$.coding[2].code') IN
('C71.0','C71.1','C71.2','C71.3','C71.4','C71.5','C71.6','C71.7','C71.8','C71.9')
      )
    GROUP BY
      json_extract_scalar(p.identifier, '$[0].value'),
      p.deceasedDateTime,
      p.birthDate
  ) base
```

```
) q
WHERE q.age_at_dx >= 18
```

**Reference SQL (Impala, HDP):**

```
WITH c71 AS (
  SELECT c_patnr, min(c_diagnose_timestamp) AS first_dx
  FROM db_tumpseudoprog_prepared.it_ishmed_diagnose
  WHERE c_diagnose_1 LIKE 'C71%' OR c_diagnose_2 LIKE 'C71%'
  GROUP BY c_patnr),
bd AS (SELECT DISTINCT c_patnr, c_birthdate FROM db_tumpseudoprog_prepared.it_ishmed_patient),
coh AS (
  SELECT c71.c_patnr, c71.first_dx,
         (year(c71.first_dx) - year(CAST(bd.c_birthdate AS TIMESTAMP))) AS age_dx
  FROM c71 JOIN bd ON c71.c_patnr = bd.c_patnr
  WHERE (year(c71.first_dx) - year(CAST(bd.c_birthdate AS TIMESTAMP))) >= 18),
death AS (SELECT DISTINCT c_patnr, c_datetimeofdeath FROM db_tumpseudoprog_prepared.it_ishmed_patient WHERE c_datetimeofdeath IS
NOT NULL)
SELECT coh.c_patnr, datediff(CAST(de.c_datetimeofdeath AS TIMESTAMP), CAST(coh.first_dx AS TIMESTAMP)) AS days_dx_to_death
FROM coh JOIN death de ON coh.c_patnr=de.c_patnr;
-- EXPORT rows; median offline. n should be ~101
```

### V16 — Deceased subgroup: sex and mean age

*Outcome (Table 2): primary thread — exact; query logged: yes. Ratings DD / GG / consensus: 1 / 1 / 1.*

**Prompt(s) as posed:**

Among the deceased adult C71 patients, what is the sex distribution and the mean age at first diagnosis? Report counts per sex and one mean age.

Platform result: F 35 / 59.5 y; M 66 / 62.2 y (n=101) | Impala reference: F 35 / 59.5 y; M 66 / 62.2 y (n=101)

**Generated SQL, primary thread — log entry Sep 01 2026 12:58 · status: data available · rows: 2**

```
SELECT
  sex,
  COUNT(DISTINCT patient_id) AS distinct_patient_count,
  AVG(age_years) AS mean_age_years
FROM (
  SELECT
    patient_id,
    sex,
    first_dx_date,
    age_years
  FROM (
    SELECT
      patient_id,
      sex,
      first_dx_date,
      age_years,
      ROW_NUMBER() OVER (PARTITION BY patient_id ORDER BY first_dx_date ASC) AS rn
    FROM (
      SELECT
        json_extract_scalar(c.subject, '$.identifier.value') AS patient_id,
        p.gender AS sex,
        date_parse(substr(MIN(c.recordedDate), 1, 10), '%Y-%m-%d') AS first_dx_date,
        date_diff(
          'year',
          date_parse(substr(p.birthDate, 1, 10), '%Y-%m-%d'),
          date_parse(substr(MIN(c.recordedDate), 1, 10), '%Y-%m-%d')
        ) AS age_years
      FROM condition c
      JOIN patient p
        ON json_extract_scalar(c.subject, '$.identifier.value') = json_extract_scalar(p.identifier, '$[0].value')
      WHERE p.deceasedDateTime IS NOT NULL
        AND p.birthDate IS NOT NULL
        AND (
              json_extract_scalar(c.code, '$.coding[0].code') IN
('C71.0','C71.1','C71.2','C71.3','C71.4','C71.5','C71.6','C71.7','C71.8','C71.9')
           OR json_extract_scalar(c.code, '$.coding[1].code') IN
('C71.0','C71.1','C71.2','C71.3','C71.4','C71.5','C71.6','C71.7','C71.8','C71.9')
           OR json_extract_scalar(c.code, '$.coding[2].code') IN
('C71.0','C71.1','C71.2','C71.3','C71.4','C71.5','C71.6','C71.7','C71.8','C71.9')
            )
      GROUP BY
        json_extract_scalar(c.subject, '$.identifier.value'),
        p.gender,
        p.birthDate
    ) extracted
    WHERE age_years >= 18
  ) ranked
```

```
  WHERE rn = 1
) final
GROUP BY sex
ORDER BY sex
```

**Reference SQL (Impala, HDP):**

```
WITH c71 AS (
  SELECT c_patnr, min(c_diagnose_timestamp) AS first_dx
  FROM db_tumpseudoprog_prepared.it_ishmed_diagnose
  WHERE c_diagnose_1 LIKE 'C71%' OR c_diagnose_2 LIKE 'C71%'
  GROUP BY c_patnr),
bd AS (SELECT DISTINCT c_patnr, c_birthdate FROM db_tumpseudoprog_prepared.it_ishmed_patient),
coh AS (
  SELECT c71.c_patnr, c71.first_dx,
         (year(c71.first_dx) - year(CAST(bd.c_birthdate AS TIMESTAMP))) AS age_dx
  FROM c71 JOIN bd ON c71.c_patnr = bd.c_patnr
  WHERE (year(c71.first_dx) - year(CAST(bd.c_birthdate AS TIMESTAMP))) >= 18),
death AS (SELECT DISTINCT c_patnr FROM db_tumpseudoprog_prepared.it_ishmed_patient WHERE c_datetimeofdeath IS NOT NULL),
g AS (SELECT DISTINCT c_patnr, c_gender FROM db_tumpseudoprog_prepared.it_ishmed_patient)
SELECT g.c_gender, count(DISTINCT coh.c_patnr) AS n, round(avg(coh.age_dx),1) AS mean_age
FROM coh JOIN death de ON coh.c_patnr=de.c_patnr JOIN g ON coh.c_patnr=g.c_patnr
GROUP BY g.c_gender;
```

#### V17 — Biopsy before resection (sequence)

*Outcome (Table 2): primary thread — not answered; query logged: —. Ratings DD / GG / consensus: 5 / 5 / 5. Re-test (new thread): exact; query logged: yes. Ratings DD / GG / consensus: 1 / 1 / 1.*

**Prompt(s) as posed:**

Among adult C71 patients who had BOTH a brain biopsy (OPS starting 1-510 or 1-511) AND a surgical resection (OPS starting 5-011 to 5-016), how many had their first biopsy strictly before their first resection? Report the count and the share of patients with both procedures.

Platform result: FAILED 2x (natural + step-by-step prompt); no answer | Impala reference: 23 / 56 (41.1%) biopsy strictly before resection

**Generated SQL, primary thread: no query logged.**

Re-test (new thread, 1 Sep 2026): platform result 23 / 56 (41.07%) exact

**Generated SQL, re-test — log entry Sep 01 2026 14:47 · status: data available · rows: 99**

```
SELECT
  q.patient_id,
  q.first_dx_date,
  q.birthDate,
  bp.first_biopsy_date,
  rs.first_resection_date
FROM (
  SELECT
    dx.patient_id,
    dx.first_dx_date,
    p.birthDate
  FROM (
    SELECT
      d.patient_id,
      MIN(d.recorded_dt) AS first_dx_date
    FROM (
      SELECT
        json_extract_scalar(c.subject, '$.identifier.value') AS patient_id,
        c.recordedDate AS recorded_dt
      FROM condition c
      WHERE
           json_extract_scalar(c.code, '$.coding[0].code') LIKE 'C71.0%'
        OR json_extract_scalar(c.code, '$.coding[0].code') LIKE 'C71.1%'
        OR json_extract_scalar(c.code, '$.coding[0].code') LIKE 'C71.2%'
        OR json_extract_scalar(c.code, '$.coding[0].code') LIKE 'C71.3%'
        OR json_extract_scalar(c.code, '$.coding[0].code') LIKE 'C71.4%'
        OR json_extract_scalar(c.code, '$.coding[0].code') LIKE 'C71.5%'
        OR json_extract_scalar(c.code, '$.coding[0].code') LIKE 'C71.6%'
        OR json_extract_scalar(c.code, '$.coding[0].code') LIKE 'C71.7%'
        OR json_extract_scalar(c.code, '$.coding[0].code') LIKE 'C71.8%'
        OR json_extract_scalar(c.code, '$.coding[0].code') LIKE 'C71.9%'
        OR json_extract_scalar(c.code, '$.coding[1].code') LIKE 'C71.0%'
        OR json_extract_scalar(c.code, '$.coding[1].code') LIKE 'C71.1%'
        OR json_extract_scalar(c.code, '$.coding[1].code') LIKE 'C71.2%'
        OR json_extract_scalar(c.code, '$.coding[1].code') LIKE 'C71.3%'
        OR json_extract_scalar(c.code, '$.coding[1].code') LIKE 'C71.4%'
        OR json_extract_scalar(c.code, '$.coding[1].code') LIKE 'C71.5%'
        OR json_extract_scalar(c.code, '$.coding[1].code') LIKE 'C71.6%'
```

```
        OR json_extract_scalar(c.code, '$.coding[1].code') LIKE 'C71.7%'
        OR json_extract_scalar(c.code, '$.coding[1].code') LIKE 'C71.8%'
        OR json_extract_scalar(c.code, '$.coding[1].code') LIKE 'C71.9%'
        OR json_extract_scalar(c.code, '$.coding[2].code') LIKE 'C71.0%'
        OR json_extract_scalar(c.code, '$.coding[2].code') LIKE 'C71.1%'
        OR json_extract_scalar(c.code, '$.coding[2].code') LIKE 'C71.2%'
        OR json_extract_scalar(c.code, '$.coding[2].code') LIKE 'C71.3%'
        OR json_extract_scalar(c.code, '$.coding[2].code') LIKE 'C71.4%'
        OR json_extract_scalar(c.code, '$.coding[2].code') LIKE 'C71.5%'
        OR json_extract_scalar(c.code, '$.coding[2].code') LIKE 'C71.6%'
        OR json_extract_scalar(c.code, '$.coding[2].code') LIKE 'C71.7%'
        OR json_extract_scalar(c.code, '$.coding[2].code') LIKE 'C71.8%'
        OR json_extract_scalar(c.code, '$.coding[2].code') LIKE 'C71.9%'
    ) d
    GROUP BY d.patient_id
  ) dx
  JOIN patient p
    ON json_extract_scalar(p.identifier, '$[0].value') = dx.patient_id
  WHERE date_diff(
          'year',
          date_parse(substr(p.birthDate, 1, 10), '%Y-%m-%d'),
          date_parse(substr(dx.first_dx_date, 1, 10), '%Y-%m-%d')
        ) >= 18
) q
JOIN (
  SELECT
    x.patient_id,
    MIN(x.proc_date) AS first_biopsy_date
  FROM (
    SELECT
      json_extract_scalar(pr.subject, '$.identifier.value') AS patient_id,
      json_extract_scalar(pr.performedPeriod, '$.start') AS proc_date
    FROM procedure pr
    WHERE
         json_extract_scalar(pr.code, '$.coding[0].code') LIKE '1-510%'
      OR json_extract_scalar(pr.code, '$.coding[1].code') LIKE '1-510%'
      OR json_extract_scalar(pr.code, '$.coding[2].code') LIKE '1-510%'
      OR json_extract_scalar(pr.code, '$.coding[0].code') LIKE '1-511%'
      OR json_extract_scalar(pr.code, '$.coding[1].code') LIKE '1-511%'
      OR json_extract_scalar(pr.code, '$.coding[2].code') LIKE '1-511%'
  ) x
  GROUP BY x.patient_id
) bp
  ON bp.patient_id = q.patient_id
JOIN (
  SELECT
    y.patient_id,
    MIN(y.proc_date) AS first_resection_date
  FROM (
    SELECT
      json_extract_scalar(pr.subject, '$.identifier.value') AS patient_id,
      json_extract_scalar(pr.performedPeriod, '$.start') AS proc_date
    FROM procedure pr
    WHERE
         json_extract_scalar(pr.code, '$.coding[0].code') LIKE '5-011%'
      OR json_extract_scalar(pr.code, '$.coding[1].code') LIKE '5-011%'
      OR json_extract_scalar(pr.code, '$.coding[2].code') LIKE '5-011%'
      OR json_extract_scalar(pr.code, '$.coding[0].code') LIKE '5-012%'
      OR json_extract_scalar(pr.code, '$.coding[1].code') LIKE '5-012%'
      OR json_extract_scalar(pr.code, '$.coding[2].code') LIKE '5-012%'
      OR json_extract_scalar(pr.code, '$.coding[0].code') LIKE '5-013%'
      OR json_extract_scalar(pr.code, '$.coding[1].code') LIKE '5-013%'
      OR json_extract_scalar(pr.code, '$.coding[2].code') LIKE '5-013%'
      OR json_extract_scalar(pr.code, '$.coding[0].code') LIKE '5-014%'
      OR json_extract_scalar(pr.code, '$.coding[1].code') LIKE '5-014%'
      OR json_extract_scalar(pr.code, '$.coding[2].code') LIKE '5-014%'
      OR json_extract_scalar(pr.code, '$.coding[0].code') LIKE '5-015%'
      OR json_extract_scalar(pr.code, '$.coding[1].code') LIKE '5-015%'
      OR json_extract_scalar(pr.code, '$.coding[2].code') LIKE '5-015%'
      OR json_extract_scalar(pr.code, '$.coding[0].code') LIKE '5-016%'
      OR json_extract_scalar(pr.code, '$.coding[1].code') LIKE '5-016%'
      OR json_extract_scalar(pr.code, '$.coding[2].code') LIKE '5-016%'
  ) y
  GROUP BY y.patient_id
) rs
  ON rs.patient_id = q.patient_id
```

##### Reference SQL (Impala, HDP):

```
WITH c71 AS (
  SELECT c_patnr, min(c_diagnose_timestamp) AS first_dx
  FROM db_tumpseudoprog_prepared.it_ishmed_diagnose
```

```
  WHERE c_diagnose_1 LIKE 'C71%' OR c_diagnose_2 LIKE 'C71%'
  GROUP BY c_patnr),
bd AS (SELECT DISTINCT c_patnr, c_birthdate FROM db_tumpseudoprog_prepared.it_ishmed_patient),
coh AS (
  SELECT c71.c_patnr, c71.first_dx,
         (year(c71.first_dx) - year(CAST(bd.c_birthdate AS TIMESTAMP))) AS age_dx
  FROM c71 JOIN bd ON c71.c_patnr = bd.c_patnr
  WHERE (year(c71.first_dx) - year(CAST(bd.c_birthdate AS TIMESTAMP))) >= 18),
bio AS (SELECT c_patnr, min(CAST(c_prozedur_begin AS TIMESTAMP)) AS first_bio FROM db_tumpseudoprog_prepared.it_ishmed_procedure
        WHERE c_prozedur_code LIKE '1-510%' OR c_prozedur_code LIKE '1-511%' GROUP BY c_patnr),
res AS (SELECT c_patnr, min(CAST(c_prozedur_begin AS TIMESTAMP)) AS first_res FROM db_tumpseudoprog_prepared.it_ishmed_procedure
        WHERE regexp_like(c_prozedur_code,'^5-01[1-6]') GROUP BY c_patnr)
SELECT count(CASE WHEN b.first_bio < r.first_res THEN 1 END) AS n_bio_before_res,
       count(*) AS n_both,
       round(100.0*count(CASE WHEN b.first_bio < r.first_res THEN 1 END)/count(*),1) AS pct
FROM coh JOIN bio b ON coh.c_patnr=b.c_patnr JOIN res r ON coh.c_patnr=r.c_patnr;
```
